# Abdominal aortic aneurysms’ histomorphology differs on the individual patient level and is not associated with classic risk factors – the HistAAA study

**DOI:** 10.1101/2024.04.16.24305904

**Authors:** Maja Carina Nackenhorst, Felix Menges, Bianca Bohmann, David Zschäpitz, Christine Bollwein, Sven Flemming, Nadja Sachs, Wolf Eilenberg, Christine Brostjan, Christoph Neumayer, Matthias Trenner, Wiebke Ibing, Hubert Schelzig, Christian Reeps, Lars Maegdefessel, Heinz Regele, Markus Udo Wagenhäuser, Claus Jürgen Scholz, Thomas Christian Gasser, Albert Busch

## Abstract

**Objective:** Abdominal aortic aneurysm (AAA) treatment is upon a diameter threshold by open (OAR) or endovascular aortic repair. So far, attempts for medical growth abrogation have failed. This study aims to elucidate the heterogeneity of AAA based on histomorphology in correlation to individual patient data and aneurysm metrics.

**Patients and Methods:** Aneurysm samples from the left anterior wall from four university center biobanks underwent histologic analysis including angiogenesis, calcification, fibrosis, type and grade of inflammation in adventitia and media. Clinical information included age, comorbidities, etc., type of aneurysm (intact, symptomatic, ruptured, inflammatory) and growth. Aneurysm morphology included diameter and semi-automated geometric analysis using Endosize^©^ (Therenva) and finite element methods (A4Clinics^©^ Research Edition, Vacops GmbH).

**Results:** 364 patients’ samples (85.4% male, median age 69 years) were evaluated and scored for acute (mixed/granulocytes) or chronic (mononuclear/plasma cells) inflammation, which was not associated with rupture (52x), symptomatic (37x; p = 0.51) or diameter (57 [52–69] mm; p = 0.87). The degree of fibrosis and the presence of angiogenesis were significantly higher (both p < 0.001) with increasing inflammation, which in turn significantly decreased with patient age (est = −0.015/year, p = 0.017). No significant differences in were seen for ruptured (vs. intact), acute (vs. elective), male (vs. female) or diabetic patients. Current smoking was associated with chronic inflammation (p = 0.007) and a higher degree of fibrosis (p = 0.03). Aneurysm geometric morphology (n=252) or annual growth rate (n=142) were not associated with histologic characteristics. Yet, local luminal thrombus formation was significantly higher with increasing inflammation (p = 0.04).

**Conclusion:** Type and degree of inflammation are the most distinguishable histologic characteristics in the AAA wall between individual patients, yet not associated with diameter or rupture. Local luminal thrombus formation is associated with inflammatory features and suggests a vivid bio-physical compartment with intra-individual differences.

## Introduction

The abdominal aortic aneurysm (AAA) is the most frequent aortic aneurysm with an age-dependent incidence of approx. 5-11% in the elderly, predominantly male population.[1, 2] While rupture is the most feared complication with high mortality rates, elective treatment is stratified on a maximum transverse diameter threshold, rapid or eccentric growth.[3, 4] Surgical exclusion by either endovascular (EVAR) or open aortic repair (OAR) is recommended by international guidelines favoring EVAR if technically feasible.

Currently, no medical treatment is available and over a dozen clinical trials on early AAA growth abrogation have failed to translate in vitro results into clinical success.[5, 6] Additionally, secondary complications such as early or late EVAR failure or suture aneurysm after OAR due to dilated sealing zones or disease progression are not infrequent.[7] Here, two shortcomings must be considered: a still incomplete understanding of the AAA pathogenesis and the possible heterogeneity of disease among individuals.

Since human tissue samples are only available from advanced disease stages, little is known about the initial and early mechanisms of AAA development.[1] A proteolytic imbalance with matrix remodeling, elastin degradation, angiogenesis and eventual calcification is believed to foster established disease.[8] Adaptive immunity might play a role in healing response.[9] The intraluminal thrombus (ILT) is considered a visco-elastic, but highly enzymatically active compartment with both, mechanical and biological properties affecting the aneurysm wall.[10, 11] Yet, the interplay of ILT, mechanical stress and biologic effect on the aneurysm wall is not understood well. A study based on a small patient series suggested a classification scheme for AAA based on pathohistologic appearances almost 30 years ago.[12] Since then, intra-individual differences have been demonstrated on expressional, biomechanical and imaging level – yet are poorly reflected in current diagnostics or clinical decision-making.[13–16] Investigations of ascending and thoracic aortic aneurysm samples have demonstrated heterogenic appearance of tissue samples within and over aneurysm entities from different sites.[17, 18] However, no such analysis is available for a sizable AAA cohort.[19]

For this study, we hypothesize a vast pathohistologic heterogeneity of AAA specimen between individual patients and aim to investigate its correlation to patient- and aneurysm-specific factors. To do so, we present a multicenter biobank based histologic evaluation of AAA samples in association with clinical data, aneurysm morphometry, growth and finite element method-based (FEM-based) identification of biomechanical factors.

### Patients and Methods

#### Patient identification, inclusion criteria, ethical statement and HistAAA consortium

The HistAAA consortium is composed of members from four university hospital vascular surgery departments with large biobanks from OAR procedures. Minimum inclusion criteria were a full thickness sample from the left anterior to midline anterior wall of the AAA sac during OAR enabling detailed histologic analysis (see below) and corresponding clinical and patient data (see below). Hence, all four biobanks from the participating institutions were screened retrospectively and the missing patient data was retrieved from electronic records. Indications for open repair were surgical reasons, patient will or operator’s choice in line with international guidelines, however, the percentage of OAR patients has declined over the study period due to more endovascular procedures.[3, 20]

All eligible patients were included consecutively. Patient data was pseudonymized for biobanking and anonymized for further analysis. The study was performed in accordance with the declaration of Helsinki and tissue sampling was approved by the local ethics committees of the individual centers (*Ethikkommission Klinikum rechts der Isar*: 2799/10; *Ethikkommission Düsseldorf:* 2019-578; *Ethikkommission Würzburg: 188/11; Ethikkommission Wien:* Ref 1729/2014). The HistAAA study was specifically approved for the leading center (*Ethikkommission Klinikum rechts der Isar*: 576/18S). Collection times were: Munich Vascular Biobank – 2005-2019; Würzburg Biobank – 2011-2015; Vienna Vascular Biobank – 2014-2018; Düsseldorf Vascular Biobank – 2017-2019;

#### Basic patient and clinical data

Basic clinical data included: age, sex (male/female), AAA state (symptomatic, ruptured, asymptomatic, inflammatory [based on i.e. halo sign in CT-angiography][21]), maximum diameter (maximum transverse diameter applying multiplane reconstructions from 1-5mm CT-angiographies 1-14 days prior to OAR (measurements performed by board-certified vascular surgeon), AAA localization (infra-, juxta-, pararenal; infrarenal = neck length ≥ 10mm), concomitant iliac aneurysm (one/two sided; common iliac artery ≥ 25mm), co-morbidities (hypertension, diabetes, hyperlipidemia, coronary artery disease CAD, chronic obstructive pulmonary disease COPD, peripheral artery disease (PAD), smoking (current/ex/never), medication (anti-thrombocyte-aggregation, angiotensin-converting-enzyme inhibitor, statin, metformin/insulin) and laboratory results (C-reactive protein CRP, leukocyte/ thrombocyte count, serum creatinine).[3] While baseline clinical data was an inclusion criterion, additional clinical data (not available for all patients, see below) was used for subgroup analysis (**Suppl. Table V**).

#### Sample acquisition, preparation and digitalization

After removal from the intraoperative situs, tissue was immediately rinsed in phosphate-buffered saline for transportation into the laboratory. Here the procedure is given for the Munich Vascular biobank, however, was similarly done in all participating centers.[22] Samples were then fixed in formalin (4% PFA) for 24 hours. If necessary, decalcification on EDTA basis (Entkalker soft SOLVAGREEN®, Carl ROTH, Karlsruhe, Germany) was performed for 2-7 days. Afterwards specimens were prepared for paraffin embedding in standard size (40 x 28 x 6.8 mm) POM histology cassettes (Kartell, Noviglio, Italy). Sections of paraffin-embedded samples (2 µm for classic samples) were mounted on glass slides (Menzel SuperFrost, 76 x 26 x 1 mm, Fisher Scientific, Schwerte, Germany).

Hematoxylin-eosin (HE) (ethanolic eosin Y solution, Mayer’s acidic hemalum solution, Waldeck, Münster, Germany) as well as elastica van Gieson (EvG) (picrofuchsin solution after Romeis 16th edition, Weigert’s solution I after Romeis 15th edition) stainings were accomplished according to the manufacturer’s protocol. Slides were covered using Pertex (Histolab products, Askim, Sweden) as mounting medium and glass coverslips (24 x 50 mm, Engelbrecht, Edermünde, Germany).

Slides (including immunohistochemistry) were then scanned with Aperio AT2 (Leica, Wetzlar, Germany), and pictures were taken with the Aperio ImageScope software (Leica). Scanned slides were analyzed and prepared for composite figures using QuPath-0.3.2 open-source software.[23]

Samples from non-aneurysmatic aortas were available for evaluation and composite figures from previous studies.[13] These samples were not included in any quantitative or qualitative analysis.

#### Histologic Analysis

Samples were analyzed by three pathologists (MN, CB, HR) according to the following criteria: *Intima*: The updated American Heart Association (AHA) classification for atherosclerotic lesions was applied to the intima.[24] Additionally, the previously described Histological Scale of Inflammation (HISA)-Score was applied to all samples.[12]

*Media*: The media was scored according to presence or absence of calcification (0 = absence, 1 = presence), degree of inflammation (0 = no inflammation; 1 = low, 2 = intermediate, 3 = high degree of inflammation), composition of inflammatory infiltrate (1 = mainly composed of mononuclear cells, 2 = granulocytes, 3 = plasma cells or 4 = mixed infiltrate) (**Fig. 1D-G, 3E-H**), presence of neoangiogenesis (0 = absence, 1 = presence) and the percentage of intact elastic fibers (<25%, 26-50%, >50%) (**Fig. 3A-D**).

**Figure 1.**
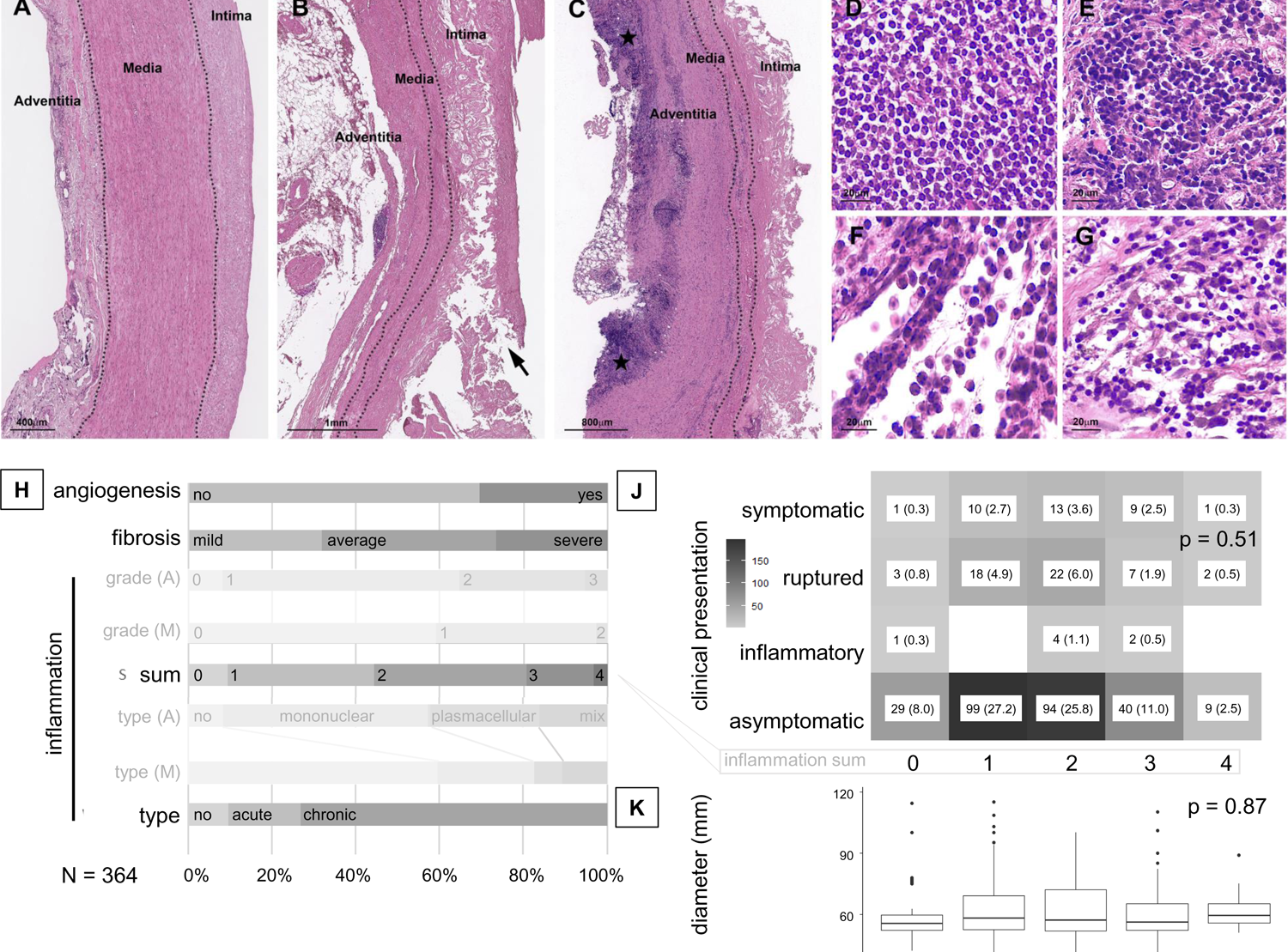
Aortic and AAA histomorphology, frequency of histologic features and correlation with clinical presentation. **(A)** Non-aneurysmatic aorta with minimal atherosclerotic changes (male patient age 67y) showing clearly distinguishable Adventitia, Media and Intima **(B)** Representative example of ruptured AAA (arrow: tear in intimal cap) with thinned media and large intimal atheroma; **(C)** Representative example of AAA with a high degree of inflammation and a thickened adventitia (asterisk mark areas with high density of inflammation) with chronic inflammation composed of mainly mononuclear cells; **(D-G)** Different types of inflammatory infiltrates: (**D,E**: chronic type) **(D)** mononuclear infiltrates; **(E)** plasma cells; (**F,G**: acute type: **(F)** neutrophils; **(G)** mixed infiltrate (all HE staining); **(H)** Percentage of selected individual histologic features and inflammation type and sum (summarized for adventitia (A) /media (M). **(J)** Frequency table for clinical AAA presentation and inflammation sum (absolute numbers, percentage, Chi-square test). **(K)** Boxplot and linear regression for AAA diameter and inflammation sum. (p < 0.05 is considered significant and highlighted bold)

*Adventitia*: Adventitial features were scored according to degree of inflammation (s. above), composition of inflammatory infiltrate (s. above) and degree of fibrosis (0 = no fibrosis, 1 = low, 2 = intermediate, 3 = high degree of fibrosis).

Degrees of inflammation and fibrosis were defined as follows:

*Inflammation*: 0 = no or only singular inflammatory cells; 1 = localized small infiltrates; 2 = localized and diffuse infiltrates; 3 = diffuse dense infiltrates (**Fig. 1B, C**).

*Fibrosis*: 0 = no proliferation of collagenous fibers; 1 = up to a third of visible adventitia with collagenous fiber proliferation; 2 = up to a half of visible adventitia with collagenous fiber proliferation; 3 = more than half of visible adventitia with collagenous fiber proliferation (**Fig. 2D-F**).

**Figure 2.**
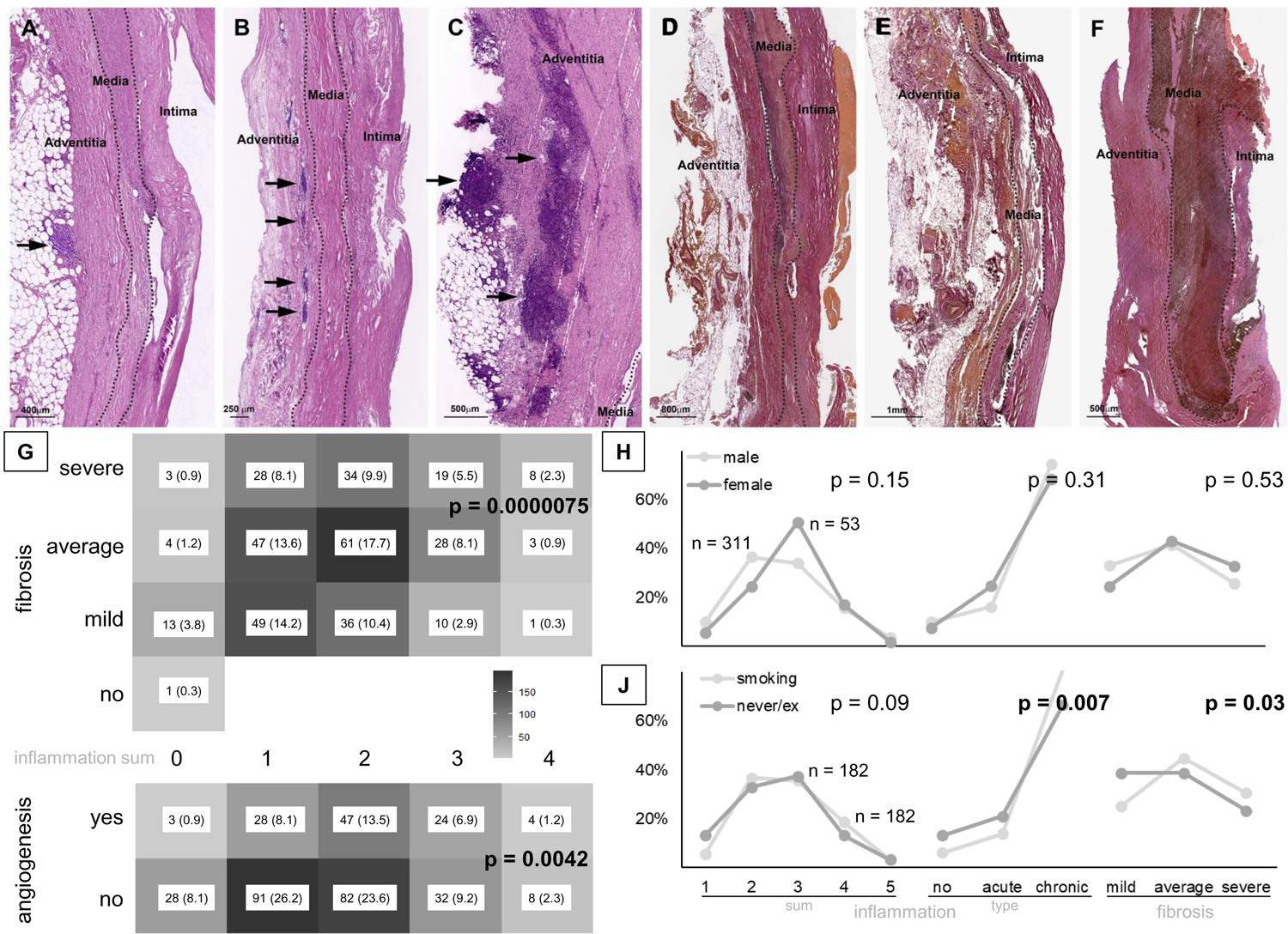
AAA fibrosis and inflammation, frequency of angiogenesis and sex comparison. **(A-C)** egrees of adventitial inflammation: **(A)** mild: only small areas of infiltrate (arrow); **(B)** average: multiple aggregates of inflammation (arrows); **(C)** severe: confluent large infiltrates (arrows) (all HE); **(D-F)** Degrees of adventitial fibrosis: **(D)** mild: <30% of adventitia; **(E)** average: 30-50%; **(F)** severe: >50% of adventitia (all EvG); **(G)** Frequency table for adventitial fibrosis (upper) or medial angiogenesis (lower) and inflammation sum, respectively (absolute numbers, percentage, Chi-square test). Relative frequencies of inflammation sum and type as well as adventitial fibrosis stratified for sex **(H)** or smoking status **(J)** (Chi-square test). (p < 0.05 is considered significant and highlighted bold)

**Figure 3.**
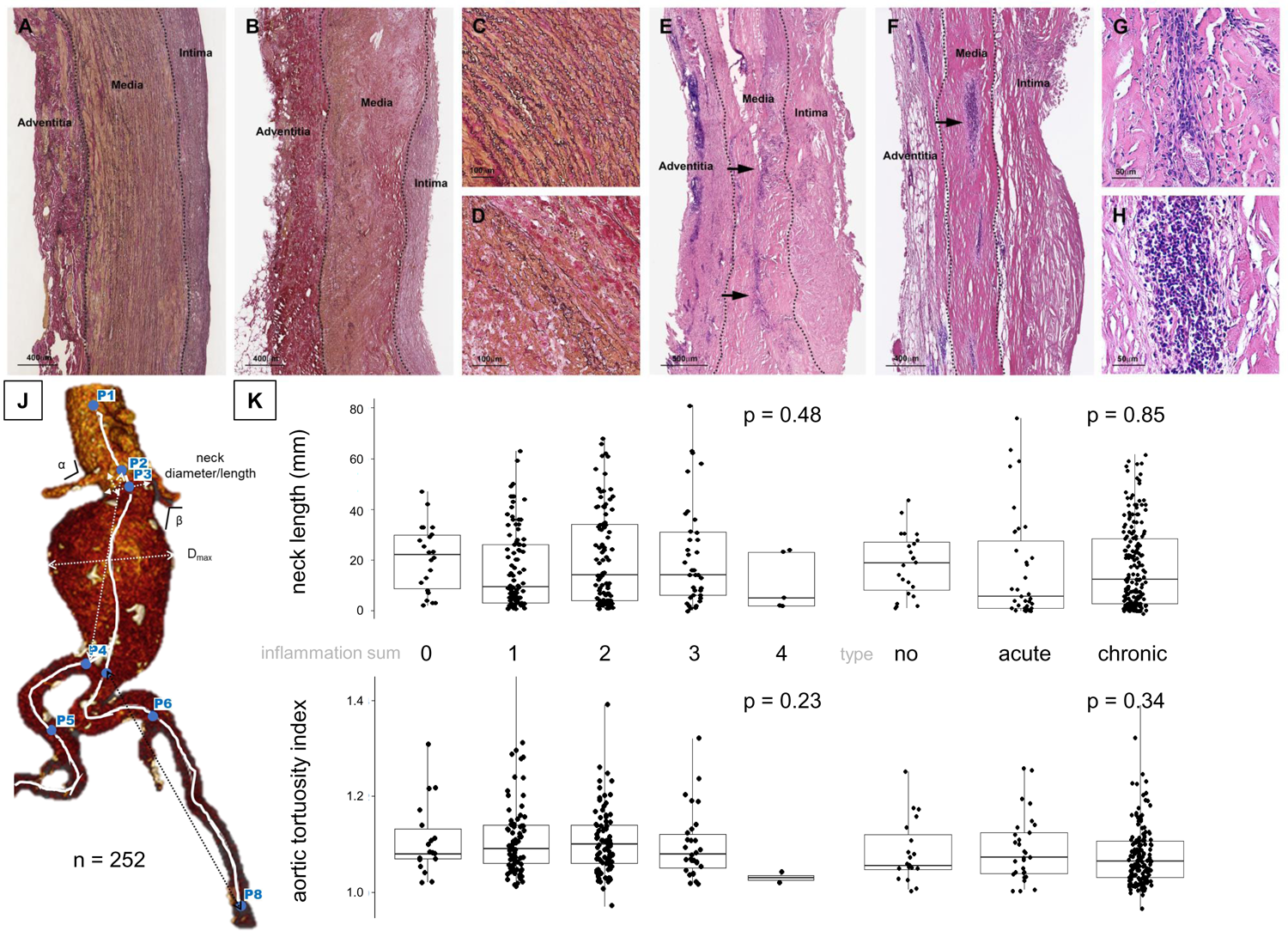
Media changes with loss of elastic fibers, inflammation and angiogenesis and AAA morphology acquisition and correlation. **(A)** Non-aneurysmatic aorta with only minimal loss/fragmentation of elastic fibers in the media; **(B)** Representative example of high degree of loss of elastic fibers in AAA (<25% intact); **(C, D)** high magnification cut outs of (A) and (B) for better visualization (all EvG). **(E, F)** Degrees of medial inflammation: **(E)** mild: focal small infiltrates (arrows); **(F)** average: various aggregates of inflammation (arrows); **(G, H)** high magnification cut-outs of (E) and (F) with perivascular infiltrates in **(G)** as a sign of medial angiogenesis (all HE). **(J)** Endosize© data acquisition: exemplary 3D CTA reconstruction after successful semi-automated segmentation using Endosize©. The neck diameter and length as well as α and β angles are calculated. The maximum AAA diameter (Dmax) is calculated perpendicular to the center line (dotted line). The aortic/iliac tortuosity indeces are calculated as the ratio centerline/raceline between P2 (lowest renal artery)/P4 (aortic bifurcation) and P4/P8 (inguinal ligament), respectively. **(K)** Boxplots with individual data points for aneurysm neck length in aortic tortuosity index in correlation to inflammation sum and type. Correlation with linear regression (sum) and ANOVA (type) (p < 0.05 is considered significant and highlighted bold)

#### Additional patient and clinical data

Additional data included height, weight and the calculated body mass index, body surface area (BSA after DuBois), aortic size index (ASI=maximum diameter/BSA).[25, 26]

*AAA growth*: Available previous CTA-based maximum transverse diameters were assessed by two vascular surgeons and included for analysis if the time difference was ≥6months. The annual growth rate was calculated by the first-last-method as described previously and extrapolated mathematically if the time difference was <12months.[27] Growth rates were then categorized as upper/lower/both middle quartiles overall and per initial diameter group (<30mm; 30-34; 35-39; 40-44; 45-49; >50mm)

#### Morphometric AAA analysis

The morphologic analysis was performed semi-automatically with Endosize^©^ (Therenva), a software for clinical assessment of AAAs as well as for EVAR planning (https://www.therenva.com/endosize) as previously described and validated by others and us.[20, 28, 29] Briefly, defined set-points were manually entered in the segmented CTA. Then, a centerline was calculated and verified with eventual manual adjustment. Calculated parameters included: suprarenal to infrarenal neck angulation (α), infrarenal neck to AAA angulation (β), maximum transverse diameter, neck length, proximal and distal neck diameter (lowest renal artery to the beginning of the aneurysm) and aortic/iliac tortuosity index (centerline to direct “raceline” distance ratio: lowest renal artery to aortic bifurcation/aortic bifurcation to inguinal ligament) (**Fig. 3J**).[30]

#### Finite element method (FEM) and local biomechanic stress estimation

A semi-automated biomechanics FEM analysis was performed using A4clinics^©^ Research Edition (Vascops GmbH).[16, 31] Briefly, a 3D model of the AAA is semi-automatically segmented from CTA images, identifying lumen, ILT and the outer contour of the vessel wall. The segmentation covers the aorta from the lowest renal artery and the bifurcation and therefore only infra/juxtarenal cases have been included. The investigators (DZ, AB) manually corrected the model if needed. A standardized arterial pressure of 140/80 mmHg was used for all FEM computations, and model output are the total vessel volume, maximal luminal/outer diameter, lumen volume, maximal ILT thickness, ILT volume, mean ILT stress, peak wall stress (PWS) and peak wall rupture index (PWRI) (**Fig. 4A**). The PWS represents the maximal stress in the wall, whilst PWRI denotes the maximum ratio between wall stress and wall strength in the aneurysm. Here, wall stress refers to the von Mises stress, a scalar representation of the 3D stress state in the vessel wall.

**Figure 4.**
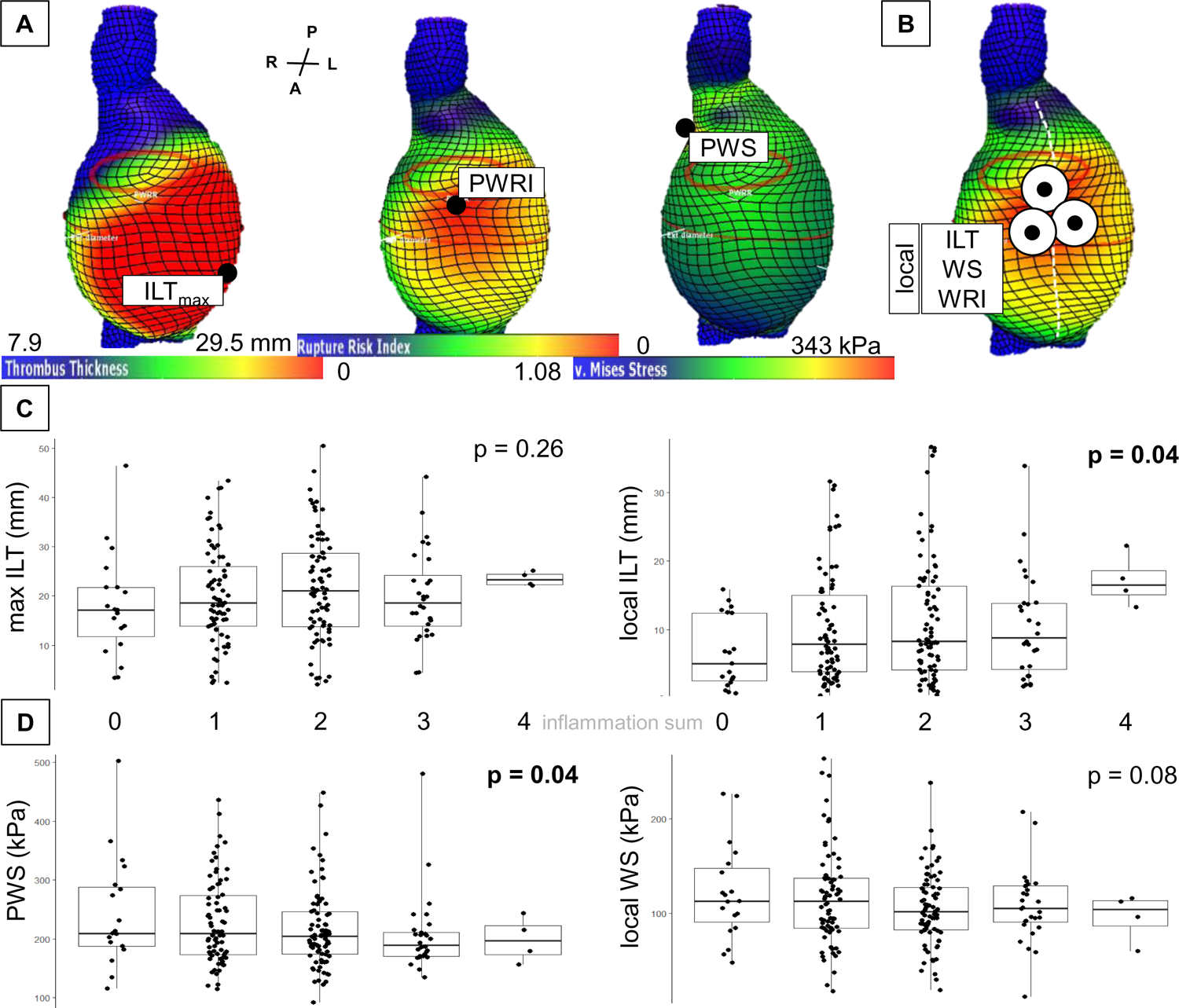
Finite element method (FEM) results (global peaks and local readings) in relation to histology findings. **(A)** FEM results displayed as heatmaps with the respective maxima: intraluminal thrombus (ILT, mm) thickness, peak wall rupture index (PWRI), and peak wall stress (PWS, kPa) marked by dots. Orientation of reconstruction represented by anterior (A), posterior (P), left (L), and right (R). **(B)** Local data acquisition by processing the mean of three readings close to the potential AAA incision line (white dotted line) during open repair. **(C)** Boxplots with individual data points for correlation of maximum and local ILT thickness with inflammation sum. **(D)** Boxplots with individual data points for correlation of peak wall stress (PWS) and local wall stress (WS) with inflammation sum. (linear regression, p < 0.05 is considered significant and highlighted bold)

In addition to the above described global AAA, the software was customized to support also a localized data analysis. The added functionality allowed the user (DZ) to pick a AAA location in the individual patient, and then statistics of geometrical and biomechanical parameters in the vicinity of 20mm of this spatial location, was provided automatically. Considering the most frequent site of wall sample acquisition, the mid or left anterior wall of the aneurysm sac, ILT thickness, wall stress (WS), and wall rupture index (WRI; local ratio between wall stress and strength) were assessed in said local analysis. Specifically, three adjacent readings were taken at 20mm distance from each other and the calculated mean was used for all correlative analysis (**Fig. 4B**).

Additionally, total, luminal and thrombus volume were measured with the software and the ratio of luminal to total AAA volume was calculated to express differences in ILT content.

### Statistics

Where applicable, average values are provided as median (interquartile range [IQR]) or mean and standard deviation. Categorical data is shown as counts and percentages. Unless indicated otherwise, continuous outcomes are analyzed by t-tests, linear regression, or analysis of variance (ANOVA). Categorical data is analyzed by chi-squared tests or logistic regression. P-values < 0.05 were considered statistically significant.

Data analysis and visualization was performed with Microsoft Excel and R version 4.0.3 (R Foundation for Statistical Computing, Vienna, Austria), the latter along with extension packages ggplot2 (visualization) and pwrss (power analysis).

## Results

### Patient cohort and histologic appearance

Overall, 364 AAA patients (85.4% male, median age 69 years) and their respective aneurysm wall sample were included. Patient characteristics are shown in **Table I**. The median diameter was 57mm [IQR: 52-69] and the majority were infra-(60.2%) and juxtarenal (28.9%) aneurysms, 52 patients presented with rupture, seven were classified as inflammatory.

**Table 1:** Patient characteristics. Values are given as absolute numbers and percentage and median with 95% confidence interval where applicable. (y=years, CAD=coronary artery disease; COPD=chronic obstructive pulmonary disease; PAOD=peripheral artery occlusive disease; renal insufficiency=serum creatinine >1.2mg/dL; ASS=aspirin; ACE=angiotensin converting enzyme; BMI = body mass index, BSA = body surface area, ASI = aortic size index; CRP=C reactive protein ↑↑ = >0.5mg/dL; leukocytes ↑↑ = count >10.000/µL, thrombocytes ↑↑ = count > 450.000/µL) (p value < 0.05 is considered significant and highlighted bold, Chi sqare/Mann Whitney U/ANOVA) (Individual data availability > 83%, for patient metrics approx. 67%)

|  |  | AAA |  |  |  |  |
| --- | --- | --- | --- | --- | --- | --- |
|  |  | total | infra- | juxta- | pararenal | p |
|  |  | 364 | 219 (60.2) | 104 (28.9) | 41 (11.3) |  |
| patient characteristics |  |  |  |  |  |  |
| male n (%) |  | 311 (85.4) | 193 (88.1) | 77 (74.0) | 41 (100.0) | <0.001 |
| age (y) |  | 69 [64-74] | 69 [63-74] | 70 [65-75] | 70 [68-74] | 0.15 |
| comorbidities | hypertension | 295 (81.0) | 174 (79.5) | 86 (82.7) | 35 (85.4) | 0.59 |
|  | diabetes | 67 (18.4) | 39 (17.8) | 19 (18.3) | 9 (22.0) | 0.11 |
|  | hyperlipidemia | 199 (54.7) | 117 (53.4) | 61 (58.7) | 21 (51.2) | 0.61 |
|  | CAD | 151 (41.5) | 81 (37.0) | 43 (41.3) | 27 (65.9) | <0.001 |
|  | COPD | 69 (19.0) | 44 (20.0) | 15 (14.4) | 10 (24.4) | 0.20 |
|  | PAOD | 68 (18.7) | 38 (17.4) | 23 (22.1) | 7 (17.1) | 0.57 |
|  | renal insufficiency | 89 (24.5) | 49 (22.4) | 22 (21.2) | 18 (43.9) | <0.001 |
| smoke | current | 182 (50.0) | 110 (50.2) | 51 (49.0) | 21 (51.2) | 0.90 |
|  | ex | 89 (24.5) | 54 (24.7) | 26 (25.0) | 9 (22.0) |  |
|  | never | 33 (9.1) | 18 (8.2) | 12 (11.5) | 3 (7.3) |  |
| medication | ASS/Clopidogrel | 241 (66.2) | 149 (68.0) | 63 (60.6) | 29 (70.7) | 0.34 |
|  | ACE inhibitor | 118 (32.4) | 69 (31.5) | 34 (32.7) | 15 (36.6) | 0.81 |
|  | statins | 184 (50.5) | 114 (52.1) | 52 (50.0) | 18 (43.9) | 0.63 |
|  | metformin | 32 (8.8) | 21 (9.6) | 6 (5.8) | 5 (12.2) | 0.38 |
|  | insulin | 11 (3.0) | 6 (2.7) | 4 (3.8) | 1 (2.4) | 0.84 |
| blood | CRP ↑↑ | 139 (38.2) | 83 (37.9) | 38 (37.5) | 17 (41.5) | 0.86 |
|  | leukocytes ↑↑ | 91 (25.0) | 55 (25.1) | 29 (27.9) | 7 (17.1) | 0.055 |
|  | thrombocytes ↑↑ | 21 (5.8) | 13 (5.9) | 7 (6.7) | 1 (2.4) | 0.60 |
| patient metrics |  |  |  |  |  |  |
| height (cm) |  | 176 [170-180] | 177 [172-180] | 174 [168-180] | 176 [172-186] | 0.025 |
| weight (kg) |  | 80 [72-93] | 84 [74-95] | 74 [68-90] | 80 [73-94] | 0.027 |
| BMI |  | 26 [24-29] | 27 [24-29] | 25 [23-29] | 26 [24-29] | 0.21 |
| BSA (m²) |  | 2.0 [1.8-2.1] | 2.0 [1.9-2.1] | 1.9 [1.8-2.1] | 2.0 [1.9-2.2] | 0.48 |
| ASI |  | 2.9 [2.6-3.5] | 2.8 [2.6-3.3] | 3.1 [2.6-3.6] | 3.0 [2.5-3.7] | 0.91 |
| aneurysm characteristics |  |  |  |  |  |  |
| ruptured |  | 52 (14.3) | 28 (12.8) | 15 (14.4) | 9 (22.0) | 0.42 |
| symptomatic |  | 37 (10.2) | 22 (10.0) | 10 (9.6) | 5 (12.2) |  |
| asymptomatic |  | 273 (75.0) | 169 (77.2) | 77 (74.0) | 27 (65.8) |  |
| inflammatory |  | 7 (1.9) | 2 (1.0) | 4 (3.8) | 1 (2.4) |  |
| D<sub>max</sub> (mm) |  | 57 [52-69] | 56 [52-65] | 57 [52-70] | 65 [56-77] | 0.0053 |
| iliac aneurysm | uni | 30 (8.2) | 20 (9.1) | 5 (4.8) | 5 (12.2) | 0.14 |
|  | bi | 25 (6.9) | 10 (4.6) | 8 (7.7) | 7 (17.1) |  |

In regard to the non-aneurysmatic aorta, histologic analysis revealed angiogenesis, fibrosis and especially type and degree of inflammation in the adventitia and the media to be distinguishable characteristics (**Fig. 1A-C/H**). Medial elastic fiber content (96.9%: <25%) or calcification (90.5%: no) varied only little (**Suppl. Table I, Suppl. Fig. 1A**). All individual histologic features could be analyzed in >95% of samples.

The type and degree of inflammation in the media and adventitia were highly significantly associated (**Fig. 1B-G**, **Suppl. Fig. 1B/C, Suppl. Table I**). Specifically, adventitial low inflammatory grade was likely to have a low medial inflammatory grade. Thus, for further analysis, the grade of inflammation was summarized for both layers. Similarly, the type of inflammation was summarized as acute (mixed infiltrate + granulocytes) or chronic (mononuclear cells + plasma cells) type and was most often synchronously reflected in both layers (**Suppl. Fig. 1B, Suppl. Table I**). Inflammation sum was equally distributed among acute and chronic inflammatory types (**Suppl. Fig. 1C, Suppl. Table II**).

### Baseline clinical data correlation

Most notably, neither type nor degree of inflammation were associated with the clinical presentation as i.e. symptomatic (n = 37) or ruptured (n = 52) or AAA diameter (**Fig. 1 J/K, Suppl. Fig. 2A/B/D**). Older patients showed significantly lower degrees of inflammation (+1y: est. = −0.015, p = 0.017) and were less likely to have chronic inflammation (p = 0.0073) (**Suppl. Fig. 2A, C**).

Angiogenesis in the media (31.6% positive) correlated significantly with higher inflammation score (p = 0.004) and was more abundant in chronic infiltrates (p = 0.015) (**Fig. 2G**, **Fig. 3G, Suppl. Fig. 3A**). Similarly, fibrosis of the adventitia was more severe with chronic type (p = 0.00038) and higher inflammation score (p = 0.0000075) (**Fig. 2D-G, Suppl. Fig. 3B**). Accordingly, the degree of fibrosis was inversely correlated to increasing patient age (+1y: est.= −2.0; p = 0.0004) and showed no association with AAA diameter (**Suppl. Fig. 3C**).

### Influence of sex, smoking and diabetes

The 53 female AAA patients (14.6%) in this study were significantly older (72 [67–77] years, p = 0.004) and had smaller aneurysm diameters at the time of operation (54 [49–60] mm, p = 0.005) (**Suppl. Table III**). Body metrics were significantly lower than in males and presentation with rupture was more frequent (26.4% vs. 12.2%. p = 0.03). Yet, no significant differences between sexes were found for the individual or summarized histologic characteristics (**Suppl. Table III, Fig. 2H**).

Interestingly, current smoking (compared to never-/ex-smokers) showed a significant trend towards more fibrosis (p=0.027) and chronic inflammation (p=0.007) (**Suppl. Table IV, Figure 2J**). Of note, no differences in histomorphology were seen for diabetic patients, and specifically when administered metformin (**Suppl. Table IV, Suppl. Fig. 5A**, data not shown).

Regarding acute (ruptured + symptomatic) cases – patient age, aneurysm diameter and percentage of females were significantly increased compared to elective cases (**Suppl. Table IV**). Yet, no differences for angiogenesis, fibrosis and inflammation score or type were seen (**Suppl. Fig. 5A, Suppl. Table IV**).

#### Subgroup analysis on morphometry and growth

Besides AAA diameter, aneurysm neck configuration and vessel tortuosity was assessed semi-automatically as described above (**Fig. 3J**).[20] This data was available from 252 patients (69.2%) (**Suppl. Table V**). Again, no significant correlation between pathohistology and aneurysm neck diameter and length or aortic and iliac tortuosity indices were seen (**Fig. 3K, Supp.** Fig. 4A, **Table I**).

Data on aneurysm growth was available from 142 patients (39%). The median annual growth rate in this cohort was 3.6 mm/y [2.5 – 5.3] (**Suppl. Table V**). No significant association between inflammation sum and type was found for the upper or lower quartile of annual growth rates for the entire cohort or based on the individual start diameter (**Suppl. Fig. 4B, C**). Additionally, no such association was seen for the upper or lower 10% of growth rates (data not shown).

#### Subgroup analysis with finite element biomechanics

A4clinicsRE analysis was done in 206 patients’ CTAs (56.6%) and delivered, besides others, the ILT thickness, the wall stress and the wall rupture index all over the AAA. Besides their global peaks (ILT_max_, PWS and PWRI) these parameters have also been explored locally at the site where the incision during open repair was made). The local ILT (10.5 ± 8.3mm) was thicker in samples with higher inflammatory scores (p = 0.04) and acute inflammation type (p = 0.02) (**Fig. 4A-C, Suppl. Fig. 5B**). No significant correlations with the general ILT thickness were observed.

Accordingly, the ration of lumen volume to total AAA volume (lumen:total volume) possibly tended to decrease with the inflammation score (p = 0.09) or type (p = 0.02), while in concordance to diameter, aneurysm volume showed no association to histologic features (**Suppl. Fig. 6B**).

Interestingly, PWS (223.3 ± 71.1 kPa) decreased with higher inflammation sum (p = 0.04), while for the local wall stress (111.9 ± 44.1 kPa), only a non-significant trend was observed (p = 0.08) (**Fig. 4D, Suppl. Fig. 5C**). No changes or significant correlations for PWRI in regards to the inflammatory characteristics were seen (**Suppl. Fig. 6C**).

## Discussion

This study demonstrates for the first time that the individual type and degree of inflammation in the aortic wall of AAA patients ranges widely between individuals. Extent of fibrosis and presence of angiogenesis are, unsurprisingly, highly significantly associated with inflammation. Most notably, no clear association of these inflammatory characteristics with aneurysm diameter were seen, while patient age was associate with less inflammatory activity. Additionally, ruptured or acute AAAs did not show any differences to intact aneurysms regarding their left ventral wall characteristics. However, acute inflammatory infiltrates and higher inflammatory cell counts were significantly associated with thicker local luminal thrombus and possibly reduced biomechanical wall stress.

Based on an à posteriori power analysis, given the current distribution of individual pathohistologic features, more than 110.000 patient samples would be required to see a significant association of inflammation sum and AAA diameter, however, only about 1000 patients for a possibly significant association to acute (rupture + symptomatic) vs. elective state of AAA (**Suppl. Fig. 7A, B**). Similarly, while the type of infiltrate was not associated with diameter, increasing the sample size by 1.4-2.0 fold would result in an expected significant difference for acute cases (**Suppl. Fig. 7C, D**).

This emphasizes previous speculations on distinct inflammatory activity and angiogenesis in ruptured AAAs, which were, however, based on small sample numbers directly taken from the potential rupture site.[32, 33] Taking into account our finding that differing annual growth rates were not reflected by a differing histology, we carefully conclude that rupture might not be the consequence of an increasing inflammatory activity of the entire aneurysm sac, yet could be associated with specific types of infiltrates.

Regarding patient age, our study demonstrated lower inflammatory scores in older patients with corresponding lack of fibrosis and angiogenesis, potentially suggesting a reduced “targeting ability” for future non-surgical treatments due to reduced “cellular activity”. Thus medical growth abrogation as suggested and partially tested by others and us, might only be applicable to specific patient populations.[5, 34, 35]

Vice versa, smoking cessation, as suggested by international guidelines, associated with reduced aneurysm growth was found to be linked to more chronic inflammation and more fibrosis suggestive of higher inflammatory activity.[1, 3, 4, 36] Targeting specific subtypes of inflammation in AAA is shaping up to be an interesting field of future research, especially in regards to targeted therapies.[34, 37, 38]

Based on a similar pathohistologic approach, Le Bruijn et al. investigated 72 AAA samples using histologic criteria, such as mesenchymal cell loss, fibrosis, transmural lymphoid infiltrates, ILT organization or micro-calcifications and concluded that abdominal and thoracic aneurysm samples show distinct differences in these categories.[19] Therefore, these criteria were included in a consensus statement on surgical pathology of the aorta from the Society for Cardiovascular Pathology and the Association for European Cardiovascular Pathology in 2016. The caveat is that they stem from investigations of the ascending aorta only and focus on the differences detected in non-classical (genetic) variants of the disease, known to make up a relevant part of thoracic aneurysm disease.[17, 39] In line with our findings, they found calcification and elastic fiber degradation varying to a lower extent, while i.e. fibrosis or angiogenesis were markedly different between individuals.[19] Regarding inflammation, Rijbroek et al. investigated the distribution of 130 AAA samples on a five item scale, also taking into account chronic or acute inflammatory infiltrates.[12] In accordance with our results, they reported lower inflammatory grades and chronic infiltrates to be predominant and also did not identify a correlation with aneurysm diameter or rupture, while younger patients showed higher inflammation scores. Their “histologic inflammation scale of aneurysm” (HISA) showed a highly significant correlation to inflammation sum in our study (p < 0.001; **Suppl. Table VI**). In line with previous results, typical atherosclerotic classification via the AHA score showed only moderate variation and did not correlate well with inflammation sum (p = 0.06; **Suppl. Table III, VI**).[19]

The seven clinically identified inflammatory aneurysms (i.e. halo sign upon CTA) included in the study showed an average inflammation sum score of 2.3±0.7 and fibrosis grade of 2.6±0.5, all with acute type inflammation (data not shown).[21, 40] As previously reported in a subset of patients, these did not correspond to the few cases of immunoglobulin G4 positive samples.[41] The high numbers of lymphoid follicles in AAA tissue reported previously were not seen in our samples.[8, 19, 42] Interestingly, distinct histomorphologies between thoracic and abdominal aneurysms become more and more obvious.[12, 17, 19, 39] This might indicate distinct disease development and morphology to be rather dependent on the embryologic origin of the aortic segment, rather than the extent of disease.[43, 44]

While aneurysm geometrics demonstrated no significant associations with histologic characteristics, the extent of ILT did and the question remains, if the thrombus influences the aortic wall or vice versa.[10] Due to its layered cell- and cytokine-rich composition, previous reports speculated on a severe influence of both mechanically and enzymatically on the structural wall properties.[11, 45] Specifically, the possible implications on PWS and PWRI by i.e. finite element analysis warrant further research.[16, 46] Here, we show that PWS decreases with inflammatory activity. In this context, experimental radiology using new tracers or specific magnetic resonance imaging probes have shown an unequal distribution of their respective target in human aortic aneurysm, both in circumferential and longitudinal direction.[47, 48]

Limitations of the study include the localization of sample acquisition. Our group previously demonstrated an uneven distribution around the circumference of the aneurysm wall in a series of five patients.[49] Here, samples were taken from the (left) anterior wall only, in line with previous studies.[12, 19] Naturally, type II error cannot be ruled out, however, might be small given the 364 patient samples provided. All results are by association only and no direct mechanism can be deducted. Thus any conclusion drawn must be considered with care. Reporting standards on AAA histology should be included in international grading schemes to enable better comparability of experimental results.[39]

## Conclusion

Inflammatory type and grade are the most distinguishable histologic characteristics in the AAA wall between individual patients. Angiogenesis and fibrosis are highly significantly associated with inflammation. No significant correlation to any patient or aneurysm specific feature, especially regarding diameter or rupture was seen. Yet, the local ILT formation increased and PWS decreased with higher inflammatory activity. This intra-individual disease heterogeneity should be considered in future aneurysm research and emphasizes the complex ever changing physico-biological properties of AAA, possibly pointing towards a decreasing biological responsiveness and thus mechanical threat in the ageing patient.

## Data Availability

All data produced in the present work are contained in the manuscript

## Acknowledgement

We are most thankful for the skilled technical assistance of Renate Hegenloh and Nadja Glucka. Simon Weidle helped with clinical data acquisition.

## Author contribution statement

MN and AB conducted the study and take overall responsibility for all steps; MN, FM, BB, DZ, SF, NS, WE, CBR, CR, LM, HR, CJS, CTG and MW collected and analyzed data; BB and CJS conducted statistical analysis; MN, CB and AB wrote the manuscript. AB acquired funding. All authors have read and approved the final version of the manuscript.

