## Supplement for "Abdominal aortic aneurysms’ histomorphology differs on the individual patient level and is not associated with classic risk factors – the HistAAA study"

#### Supplement Figures

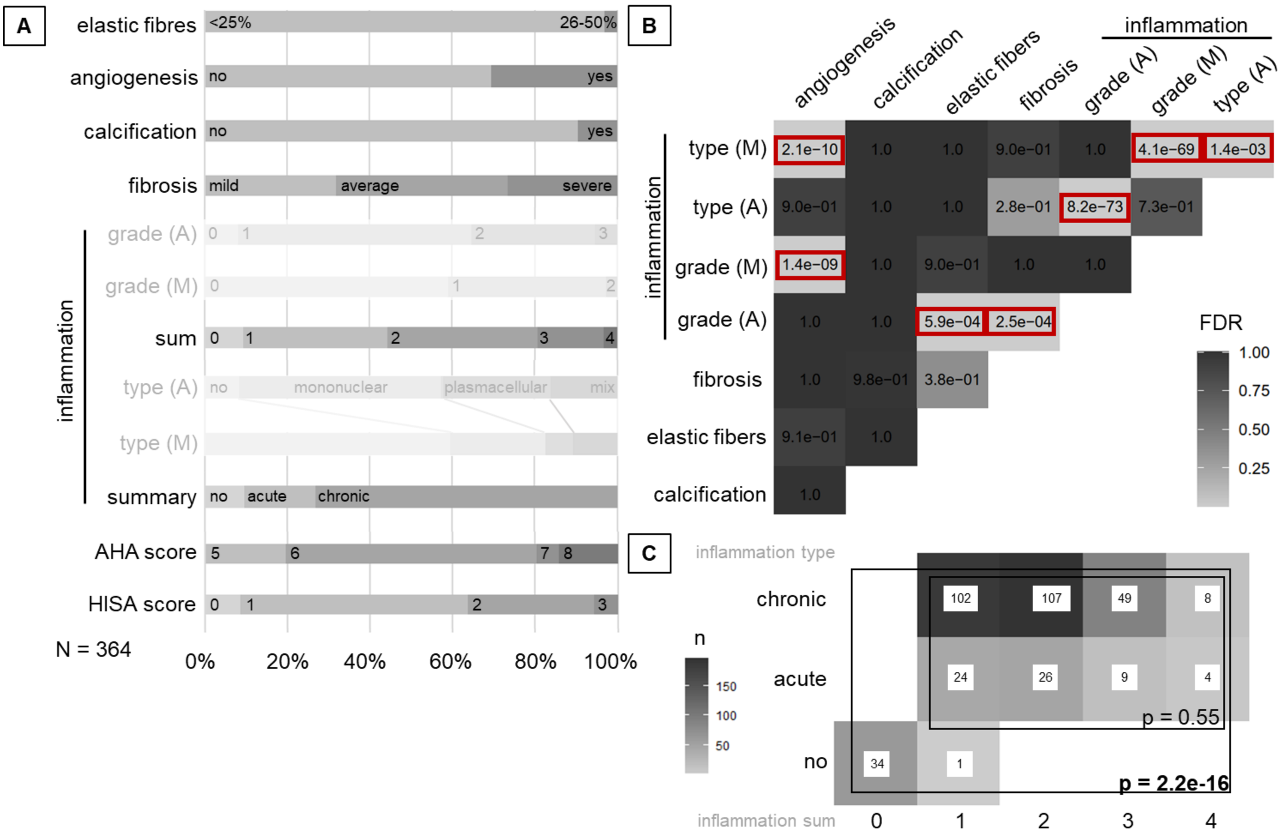

**Suppl. Figure 1. (A)** Percentage of all individual histologic features and inflammation sum and score (summarized for adventitia (A) /media (M)). **(B)** Pairwise comparison of all variables displaying false discovery rates (FDRs) from chi-squared tests; FDRs<0.05 are highlighted. **(C)** Frequency table for type and inflammation sum (A+M) (absolute numbers; Chi-square test). (p < 0.05 is considered significant and highlighted bold or marked red)

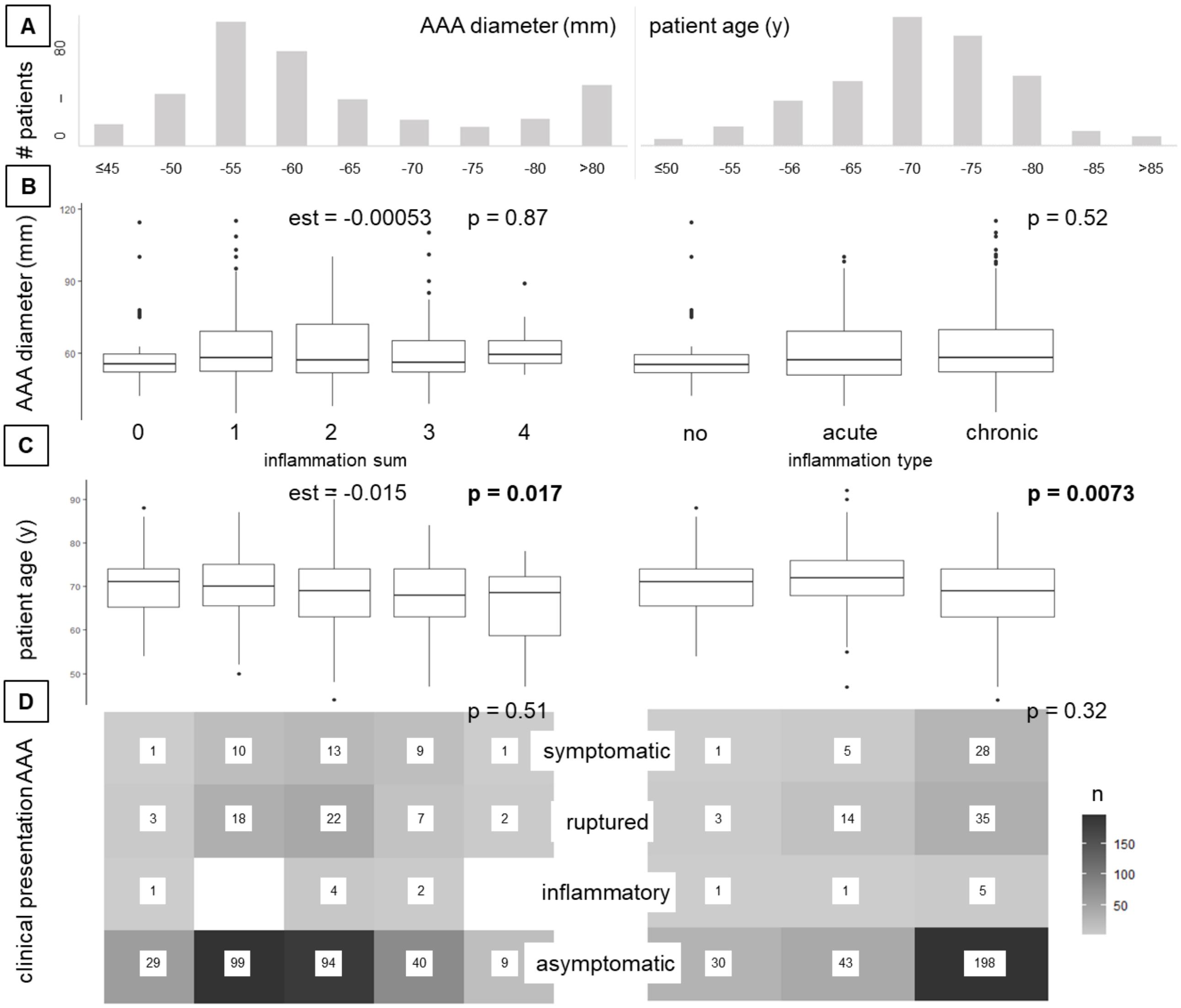

**Suppl. Figure 2. (A)** Frequency of AAA diameter and patient age in the study cohort. **(B)** Boxplot and linear regression/ANOVA for AAA diameter with inflammation sum and type. **(C)** Boxplot and linear regression/ANOVA for patient age with inflammation sum and type. **(D)** Frequency table for clinical AAA presentation with inflammation sum and type (absolute numbers, Chi-square test). ($p < 0.05$ is considered significant and highlighted bold)

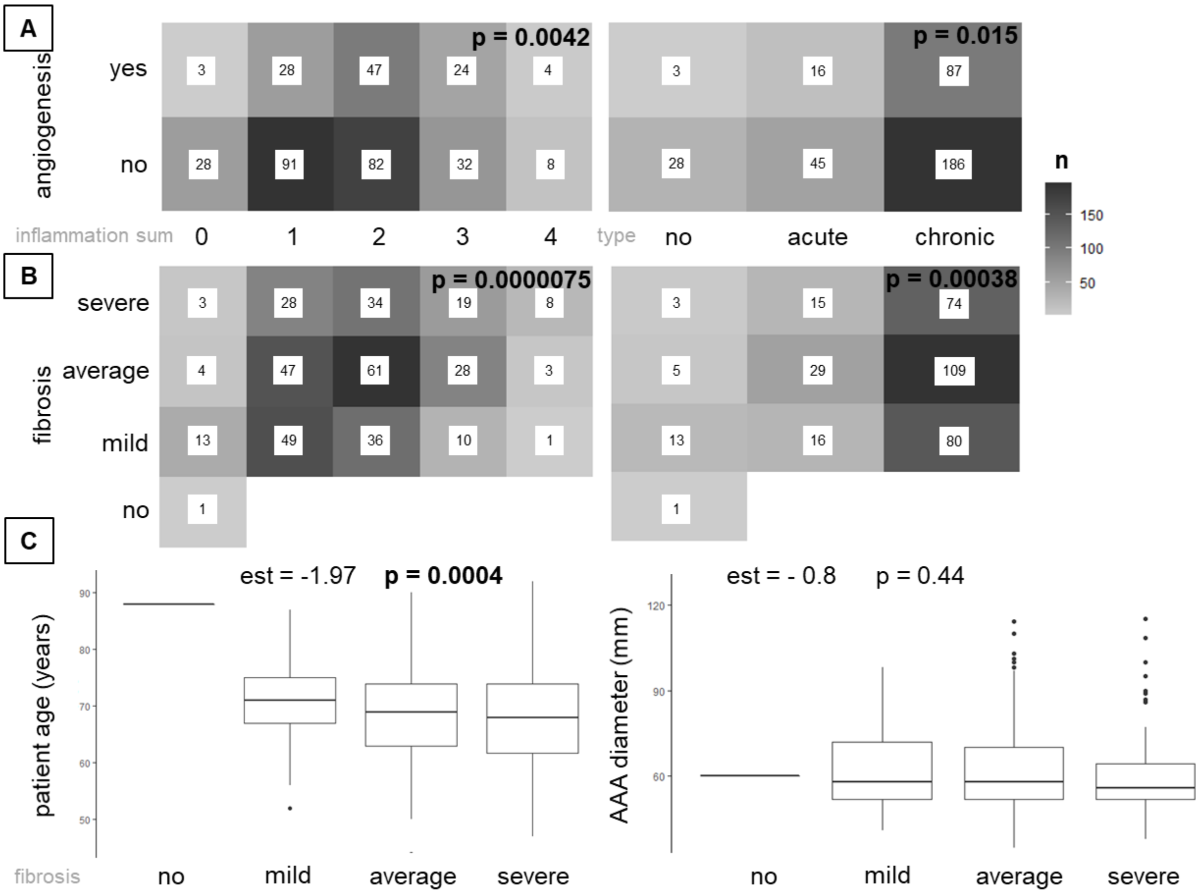

**Suppl. Figure 3. (A)** Frequency of angiogenesis with inflammation sum or type (absolute numbers; Chi-square test). **(B)** Frequency table for fibrosis with inflammation score and type (absolute numbers, Chi-square test). Boxplots and linear regression for patient age and AAA diameter with fibrosis (est. = estimate). ($p < 0.05$ is considered significant and highlighted bold)

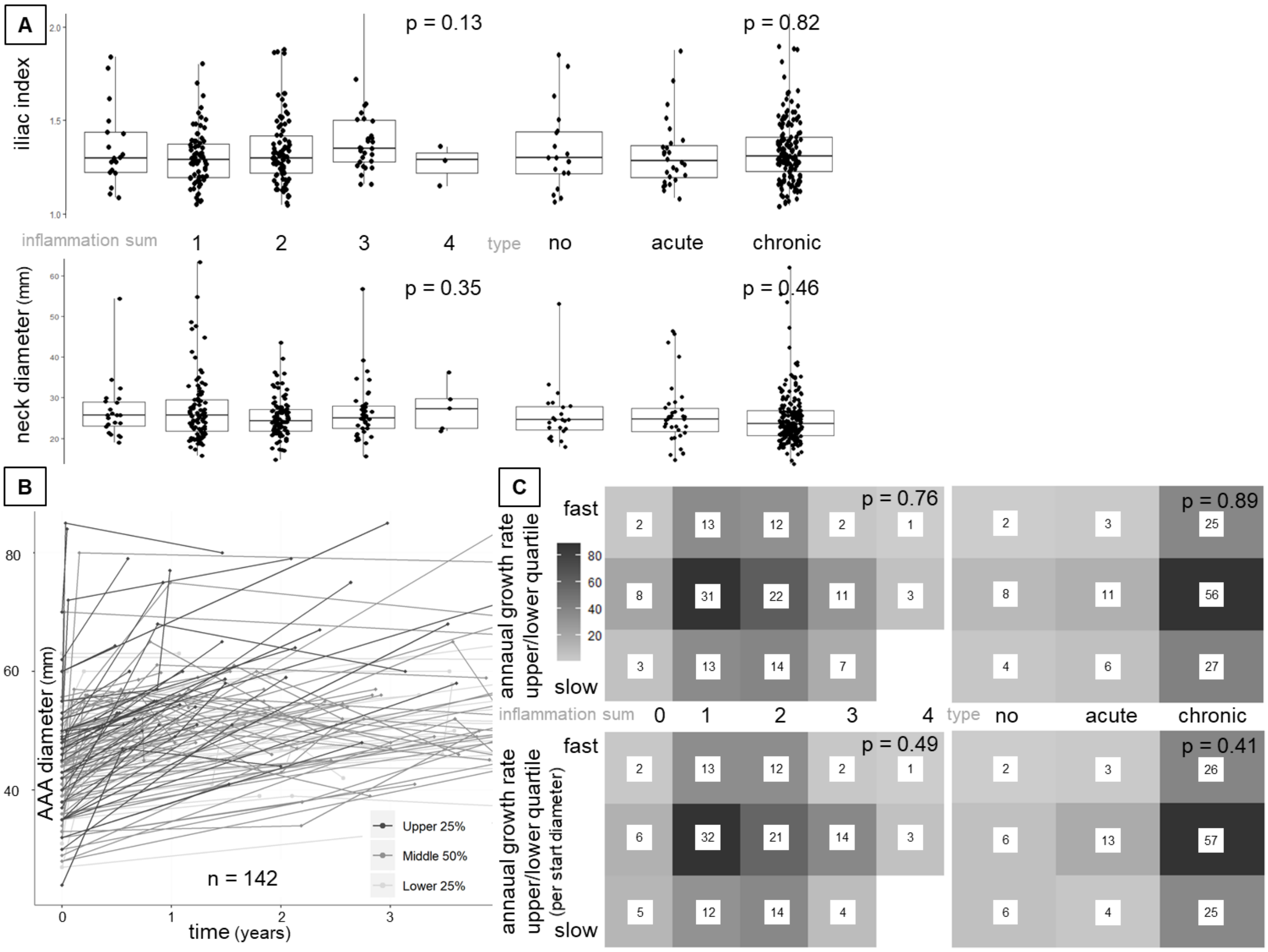

**Suppl. Figure 4. (A)** Boxplots with individual data points for aneurysm neck diameter and iliac tortuosity index in correlation to inflammation sum and type. Correlation with linear regression (sum) and ANOVA (type). **(B)** Individual aneurysm growth sorted by upper and lower quartile for the 142 patients with available growth data. **(C)** Frequency table for aneurysm growth and inflammation sum and type, respectively (absolute numbers, Chi-square test). Shown are the overall cohorts' upper/2 middle/lower quartile (upper panel) and sorted quartiles by initial diameter (lower panel). ($p < 0.05$ is considered significant and highlighted bold)

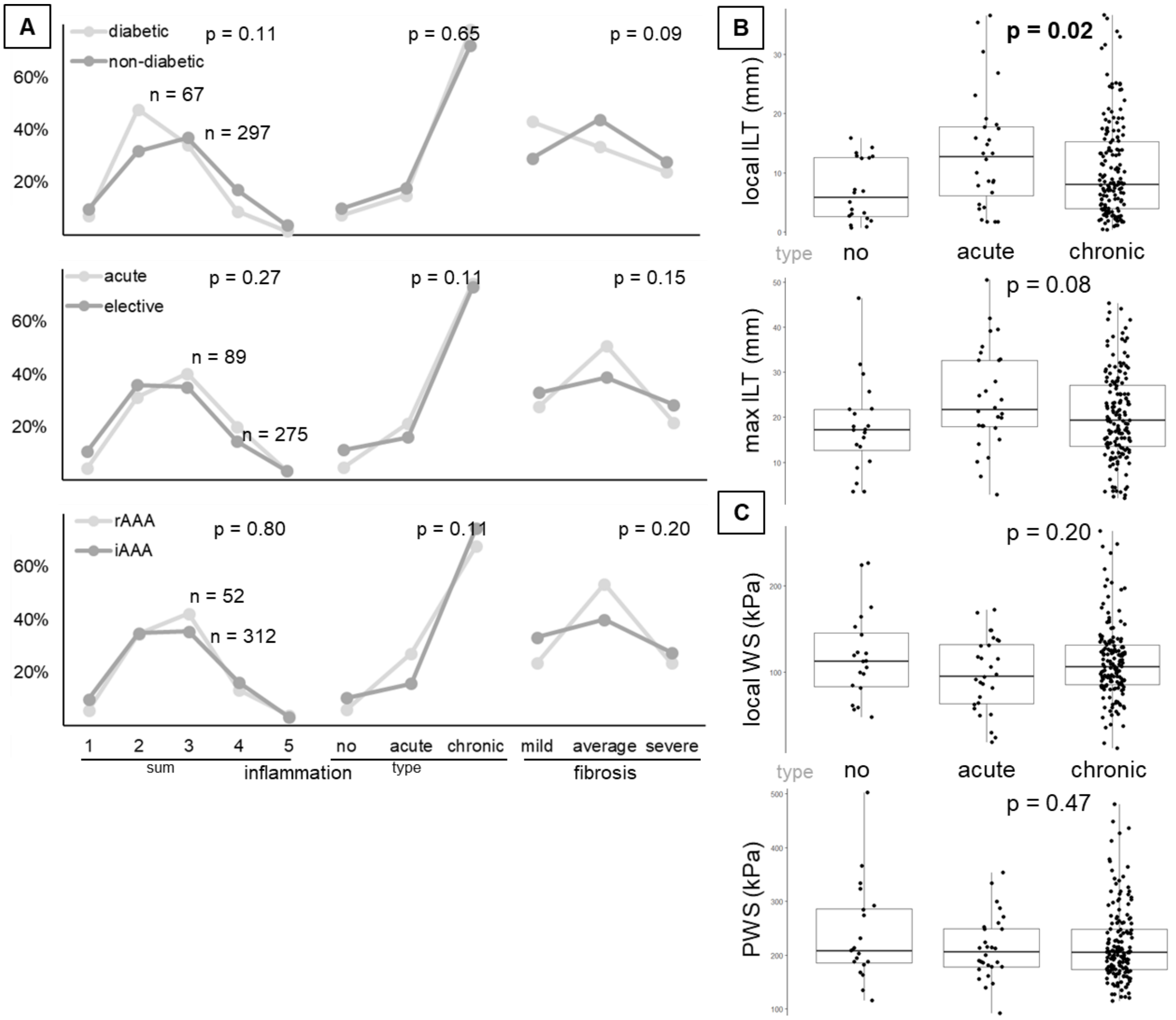

**Suppl. Figure 5. (A)** Relative frequencies of inflammation sum and type and adventitial fibrosis stratified for diabetes vs. non-diabetic, acute vs. elective operation and ruptured vs. intact AAA (Chi-square test). **(B)** Boxplots with individual data points for maximum and local ILT in correlation to inflammation type (ANOVA). **(C)** Boxplots with individual data points for peak wall stress (PWS) and local wall stress (WS) in correlation to inflammation type (ANOVA). ($p < 0.05$ is considered significant and highlighted bold)

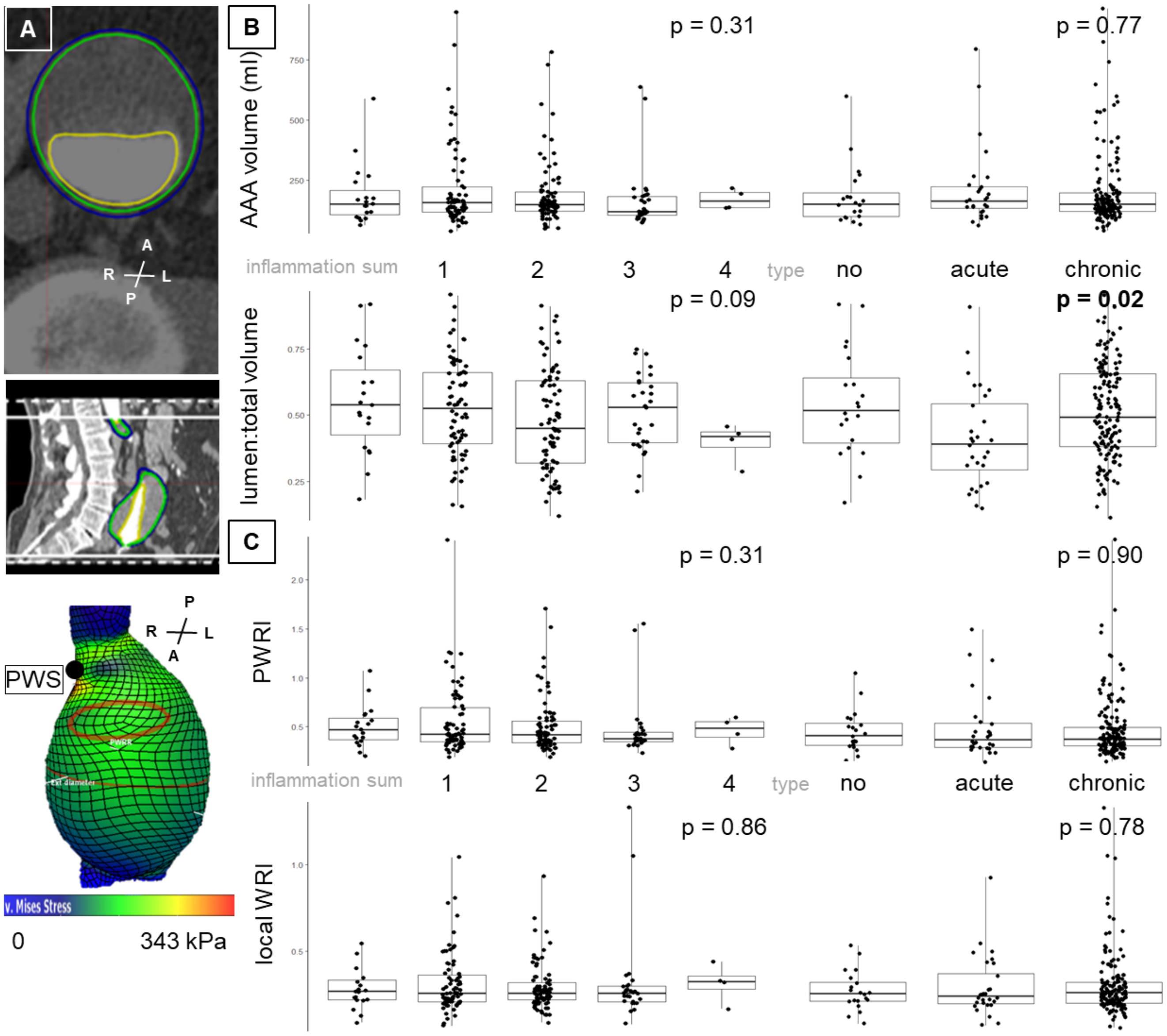

**Suppl. Figure 6. (A)** Finite element method-based (FEM-based) post-processing of CTA images. Wall stress (kPa) displayed as heatmap with its peak (peak wall stress; PWS) indicated by the dot. Orientation of reconstruction represented by anterior (A), posterior (P), left (L), and right (R). **(B)** Boxplots with individual data points for aneurysm volume and the ratio of luminal to total volume (lumen:total volume) in relation to inflammation sum and type. Correlation with linear regression (sum) and ANOVA (type). **(C)** Boxplots with individual data points for peak wall rupture index (PWRI) and local wall rupture index (WRI) in relation to inflammation sum and type. Correlation with linear regression (sum) and ANOVA (type). ($p < 0.05$ is considered significant and highlighted bold)

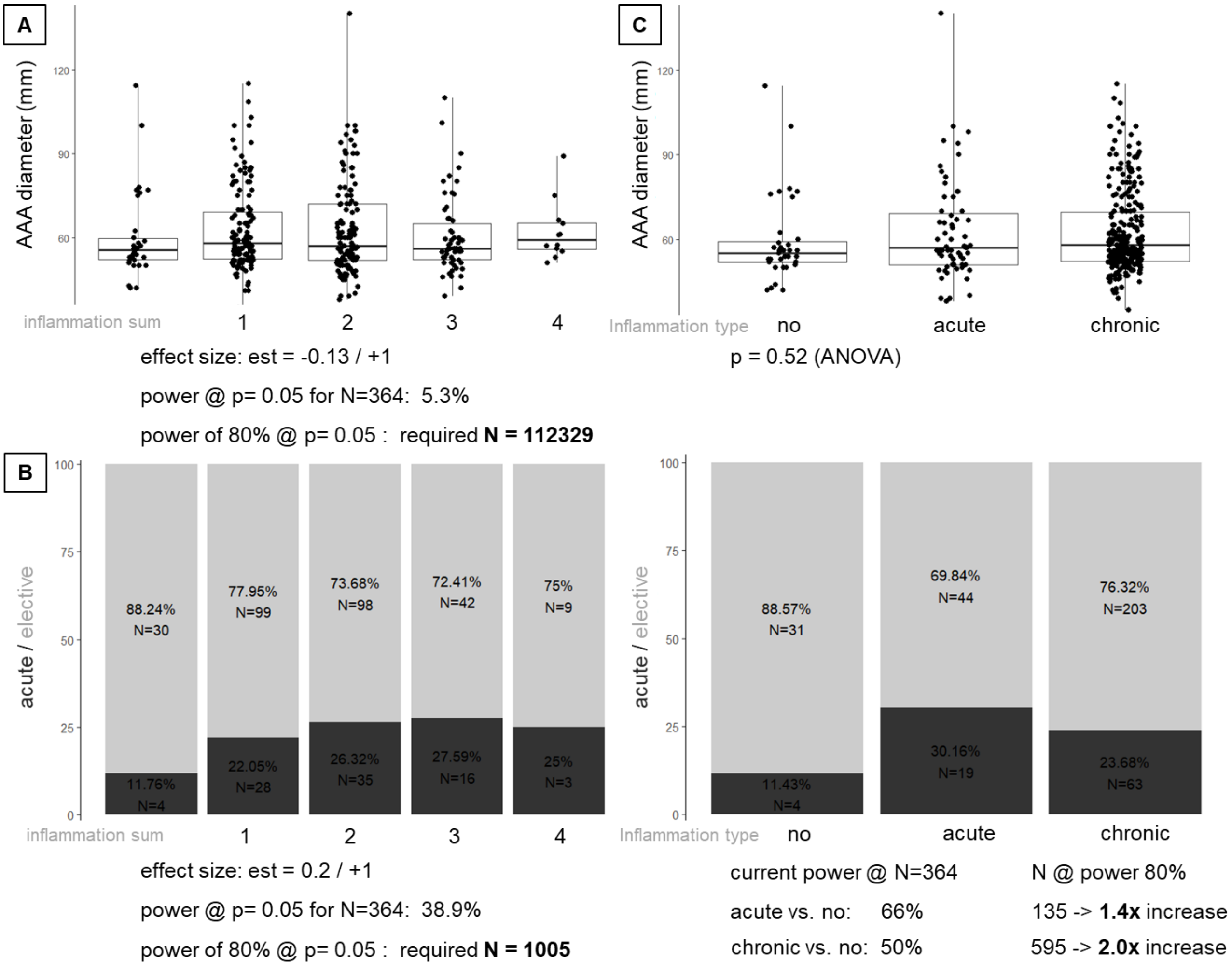

**Suppl. Figure 7. A posteriori power analysis. (A)** Boxplots with individual data points for aneurysm diameter in correlation to inflammation sum with linear regression analysis, corresponding power analysis and sample size estimation for reaching 80% power. **(B)** Proportion graph acute vs. elective cases in correlation to inflammation sum with effect size (logistic regression), corresponding power analysis and sample size estimation for reaching 80% power. **(C)** Boxplots with individual data points for aneurysm diameter in correlation to inflammation type. (ANOVA for group differences, p < 0.05 is considered significant and highlighted bold) **(D)** Proportion graph acute vs. elective cases in correlation to inflammation type with effect size (logistic regression), corresponding power analysis and sample size estimation for reaching 80% power.

### Supplement Tables

| | | | | media | | | | | | | | | | | | | | | | adventitia | | | | | | | | | | | |
|---|---|---|---|---|---|---|---|---|---|---|---|---|---|---|---|---|---|---|---|---|---|---|---|---|---|---|---|---|---|---|---|
| | | | | calc | | inflammation | | | | | | | | | | angio | | fibers | | inflammation | | | | | | | | | fibrosis | | |
| | | | | | | grade | | | | type | | | | | | | | | | grade | | | | type | | | | | | | |
| | | | | n | y | no | mild | average | severee | no | monocytes | granulocytes | plasma cells | mix | no | yes | <25% | <50% | no | mild | average | severee | no | monocytes | granulocytes | plasma cells | mix | no | mild | average | severee |
| media | calc | | n | 315 | | | | | | | | | | | | | | | | | | | | | | | | | | | |
| media | calc | | y | | 33 | | | | | | | | | | | | | | | | | | | | | | | | | | |
| media | inflammation | grade | 0 | 181 | 23 | 204 | | | | | | | | | | | | | | | | | | | | | | | | | |
| media | inflammation | grade | 1 | 123 | 9 | | 132 | | | | | | | | | | | | | | | | | | | | | | | | |
| media | inflammation | grade | 2 | 8 | 1 | | | 9 | | | | | | | | | | | | | | | | | | | | | | | |
| media | inflammation | grade | 3 | 0 | 0 | | | | 0 | | | | | | | | | | | | | | | | | | | | | | |
| media | inflammation | type | 0 | 182 | 24 | 204 | 1 | 0 | 0 | 205 | | | | | | | | | | | | | | | | | | | | | |
| media | inflammation | type | 1 | 75 | 4 | 0 | 74 | 5 | 0 | | 79 | | | | | | | | | | | | | | | | | | | | |
| media | inflammation | type | 2 | 0 | 0 | 0 | 0 | 0 | 0 | | | 0 | | | | | | | | | | | | | | | | | | | |
| media | inflammation | type | 3 | 23 | 0 | 0 | 22 | 1 | 0 | | | | 23 | | | | | | | | | | | | | | | | | | |
| media | inflammation | type | 4 | 32 | 5 | 0 | 34 | 3 | 0 | | | | | 37 | | | | | | | | | | | | | | | | | |
| media | angio | | n | 117 | 24 | 170 | 64 | 6 | 0 | 171 | 36 | 0 | 8 | 24 | 241 | | | | | | | | | | | | | | | | |
| media | angio | | y | 97 | 9 | 33 | 68 | 3 | 0 | 33 | 43 | 0 | 15 | 13 | | 106 | | | | | | | | | | | | | | | |
| media | fibers | | 0 | 209 | 30 | 190 | 130 | 9 | 0 | 190 | 81 | 0 | 23 | 37 | 224 | 106 | 338 | | | | | | | | | | | | | | |
| media | fibers | | 1 | 9 | 2 | 11 | 0 | 0 | 0 | 11 | 0 | 0 | 0 | 0 | 11 | 0 | | 11 | | | | | | | | | | | | | |
| adventitia | inflammation | grade | 0 | 25 | 2 | 20 | 7 | 0 | 0 | 20 | 3 | 0 | 0 | 4 | 22 | 5 | 24 | 4 | 29 | | | | | | | | | | | | |
| adventitia | inflammation | grade | 1 | 167 | 23 | 112 | 72 | 4 | 0 | 113 | 45 | 0 | 16 | 15 | 124 | 66 | 185 | 4 | | 198 | | | | | | | | | | | |
| adventitia | inflammation | grade | 2 | 94 | 8 | 53 | 43 | 4 | 0 | 53 | 25 | 0 | 7 | 15 | 72 | 29 | 100 | 3 | | | 105 | | | | | | | | | | |
| adventitia | inflammation | grade | 3 | 17 | 0 | 9 | 8 | 0 | 0 | 9 | 6 | 0 | 0 | 2 | 13 | 4 | 17 | 0 | | | | 19 | | | | | | | | | |
| adventitia | inflammation | type | 0 | 25 | 2 | 20 | 7 | 0 | 0 | 20 | 3 | 0 | 0 | 4 | 22 | 5 | 24 | 4 | 29 | 0 | 0 | 0 | 29 | | | | | | | | |
| adventitia | inflammation | type | 1 | 146 | 16 | 92 | 67 | 2 | 0 | 93 | 50 | 0 | 4 | 13 | 115 | 47 | 159 | 4 | 0 | 83 | 74 | 14 | | 171 | | | | | | | |
| adventitia | inflammation | type | 2 | 2 | 0 | 1 | 1 | 0 | 0 | 1 | 1 | 0 | 0 | 0 | 1 | 1 | 2 | 0 | 0 | 2 | 0 | 0 | | | 2 | | | | | | |
| adventitia | inflammation | type | 3 | 82 | 8 | 54 | 34 | 1 | 0 | 54 | 16 | 0 | 14 | 5 | 51 | 38 | 87 | 2 | 0 | 74 | 16 | 1 | | | | 91 | | | | | |
| adventitia | inflammation | type | 4 | 47 | 7 | 27 | 22 | 5 | 0 | 27 | 9 | 0 | 5 | 13 | 41 | 13 | 53 | 4 | 0 | 37 | 15 | 4 | | | | | 56 | | | | |
| adventitia | fibrosis | | 0 | 0 | 0 | 0 | 0 | 0 | 0 | 0 | 0 | 0 | 0 | 0 | 0 | 0 | 0 | 0 | 1 | 0 | 0 | 0 | 1 | 0 | 0 | 0 | 0 | 1 | | | |
| adventitia | fibrosis | | 1 | 86 | 16 | 65 | 37 | 0 | 0 | 65 | 22 | 0 | 9 | 6 | 74 | 28 | 97 | 7 | 15 | 71 | 22 | 1 | 15 | 47 | 0 | 30 | 15 | | 109 | | |
| adventitia | fibrosis | | 2 | 106 | 10 | 76 | 57 | 3 | 0 | 77 | 35 | 0 | 12 | 12 | 85 | 51 | 139 | 0 | 5 | 81 | 50 | 7 | 5 | 71 | 1 | 37 | 28 | | | 143 | |
| adventitia | fibrosis | | 3 | 84 | 7 | 45 | 39 | 5 | 0 | 48 | 22 | 0 | 2 | 16 | 66 | 24 | 84 | 4 | 6 | 44 | 31 | 11 | 6 | 51 | 1 | 22 | 12 | | | | 92 |

**Suppl. Table I. Frequency of histologic features.** Cross table of the absolute numbers of all individual histologic characteristics for media and adventitia. (calc = calcification; angio = angiogenesis; fibers = # of elastic fibers)

| | | total | wall sample inflammatory type | | | p |
|---|---|---|---|---|---|---|
| | | | no | acute | chronic | |
| | | **364** | 35 () | 63 () | 266 () | |
| **patient characteristics** | | | | | | |
| male n (%) | | 311 (85.4) | 31 () | 50 () | 230 () | 0.28 |
| age (y) | | 69 [64-74] | 71 [66-74] | 72 [68-76] | 69 [63-74] | **0.0073** |
| comorbidities | hypertension | 295 (81.0) | | | | |
| | diabetes | 67 (18.4) | | | | |
| | hyperlipidemia | 199 (54.7) | | | | |
| | CAD | 151 (41.5) | | | | |
| | COPD | 69 (19.0) | | | | |
| | PAOD | 68 (18.7) | | | | |
| | renal insufficiency | 89 (24.5) | | | | |
| smoke | current | 182 (50.0) | | | | |
| | ex | 89 (24.5) | | | | |
| | never | 33 (9.1) | | | | |
| medication | ASS/Clopidogrel | 241 (66.2) | | | | |
| | ACE inhibitor | 118 (32.4) | | | | |
| | statins | 184 (50.5) | | | | |
| | metformin | 32 (8.8) | | | | |
| | insulin | 11 (3.0) | | | | |
| blood | CRP ↑↑ | 139 (38.2) | | | | |
| | leukocytes ↑↑ | 91 (25.0) | | | | |
| | thrombocytes ↑↑ | 21 (5.8) | | | | |
| **patient metrics** (data completeness 67 - 74 %) | | | | | | |
| height (cm) | | 176 [170-180] | 178 [170-181] | 175 [168-178] | 176 [170-181] | 0.51 |
| weight (kg) | | 80 [72-93] | 88 [77-93] | 80 [70-87] | 80 [72-94] | 0.39 |
| **aneurysm characteristics** | | | | | | |
| ruptured<br>symptomatic<br>asymptomatic<br>inflammatory | | 52 (14.3)<br>37 (10.2)<br>273 (75.0)<br>7 (1.9) | 3 ()<br>1 ()<br>30 ()<br>1 () | 14 ()<br>5 ()<br>44 ()<br>1 () | 35 ()<br>31 ()<br>199 ()<br>4 () | 0.26 |
| $D_{max}$ (mm) | | 57 [52-69] | 55 [52-59] | 57 [51-69] | 58 [52-70] | 0.53 |
| iliac aneurysm | uni | 30 (8.2) | 1 () | 1 () | 28 () | **0.028** |
| | bi | 25 (6.9) | 5 () | 3 () | 15 () | |
| **histo-morphology** | | | | | | |
| angiogenesis (yes) n (%) | | 106 (30.5) | 3 (8.6) | 16 (25.4) | 87 (32.7) | **0.015** |
| fibrosis | mild | 109 (31.6) | 14 () | 16 () | 80 () | **<0.001** |
| | average | 143 (41.4) | 5 () | 29 () | 109 () | |
| | severe | 92 (26.7) | 3 () | 15 () | 74 () | |
| inflammation sum | 0 | 34 (9.3) | 34 () | - | - | **<0.001** |
| | 1 | 127 (34.9) | 1 () | 24 () | 102 () | |
| | 2 | 133 (36.5) | - | 26 () | 107 () | |
| | 3 | 58 (15.9) | - | 9 () | 49 () | |
| | 4 | 12 (3.3) | - | 4 () | 8 () | |

**Suppl. Table II: Inflammatory infiltrate characteristics.** Values are given as absolute numbers and percentage and median with 95% confidence interval where applicable. (y=years, CAD=coronary artery disease; COPD=chronic obstructive pulmonary disease; PAOD=peripheral artery occlusive disease; renal insufficiency=serum creatinine >1.2mg/dL; ASS=aspirin; ACE=angiotensin converting enzyme; BMI = body mass index, BSA = body surface area, ASI = aortic size index; CRP=C reactive protein ↑↑ = >0.5mg/dL; leukocytes ↑↑ = count >10.000/µL, thrombocytes ↑↑ = count > 450.000/µL) (p value < 0.05 is considered significant and highlighted bold, Chi sqare/Mann Whitney U/ANOVA) (Individual data availability > 83%)

| | | | total | male | female | p |
|---|---|---|---|---|---|---|
| | | | **364** | 311 | 53 | - |
| **patient characteristics** | | | | | | |
| age (y) | | | 69 [64-74] | 69 [64-74] | 72 [67-77] | **0.0041** |
| comorbidities | hypertension | | 295 (81.0) | 259 (83.3) | 37 (69.8) | **0.02** |
| | diabetes | | 67 (18.4) | 59 (19.0) | 8 (15.1) | 0.50 |
| | hyperlipidemia | | 199 (54.7) | 177 (56.9) | 22 (41.5) | **0.037** |
| | CAD | | 151 (41.5) | 134 (43.1) | 17 (32.1) | 0.13 |
| | COPD | | 69 (19.0) | 56 (18.0) | 13 (24.5) | 0.26 |
| | PAOD | | 68 (18.7) | 57 (18.3) | 11 (20.8) | 0.68 |
| | renal insufficiency | | 89 (24.5) | 79 (25.4) | 10 (18.9) | 0.31 |
| smoke | current | | 182 (50.0) | 158 (50.8) | 24 (45.3) | 0.31 |
| | ex | | 89 (24.5) | 80 (25.7) | 9 (17.0) | |
| | never | | 33 (9.1) | 27 (8.7) | 7 (13.2) | |
| medication | ASS/Clopidogrel | | 241 (66.2) | 204 (80.6) | 37 (82.7) | 0.55 |
| | ACE inhibitor | | 118 (32.4) | 103 (40.7) | 15 (33.1) | 0.49 |
| | statins | | 184 (50.5) | 162 (64.0) | 23 (51.0) | 0.24 |
| | metformin | | 32 (8.8) | 29 (11.5) | 3 (7.1) | 0.38 |
| | insulin | | 11 (3.0) | 9 (3.6) | 2 (4.5) | 0.73 |
| blood | CRP elevated | | 139 (38.2) | 119 (38.0) | 20 (37.7) | 0.94 |
| | leukocytes elevated | | 91 (25.0) | 70 (23.1) | 19 (35.8) | 0.055 |
| | thrombocytes elevated | | 21 (5.8) | 14 (5.1) | 3 (5.7) | 0.71 |
| **body metrics** | | | | | | |
| height (cm) | | | 176 [170-180] | 177 [172-182] | 166 [160-168] | **<0.001** |
| weight (kg) | | | 80 [72-93] | 85 [75-95] | 62 [54-68] | **<0.001** |
| BMI | | | 26 [24-29] | 27 [24-29] | 22 [20-25] | **<0.001** |
| BSA ($m^2$) | | | 2.0 [1.8-2.1] | 2.0 [1.88-2.14] | 1.66 [1.56-1.76] | 0.22 |
| ASI | | | 2.9 [2.6-3.5] | 2.88 [2.54-3.48] | 3.1 [2.8-3.4] | 0.83 |
| **aneurysm characteristics** | | | | | | |
| ruptured<br>symptomatic<br>asymptomatic<br>inflammatory | | | 52 (14.3)<br>37 (10.2)<br>273 (75.0)<br>7 (1.9) | 38 (12.2)<br>31 (10.0)<br>240 (77.2)<br>5 (1.6) | 14 (26.4)<br>6 (11.3)<br>33 (62.3)<br>2 (3.8) | **0.033** |
| pararenal<br>iuxtarenal<br>infrarenal | | | 41 (11.3)<br>104 (28.6)<br>219 (60.2) | 41 (13.2)<br>77 (24.8)<br>193 (62.1) | 0<br>27 (50.9)<br>26 (49.1) | **<0.001** |
| $D_{max}$ (mm) | | | 57 [52-69] | 58 [53-70] | 54 [49-60] | **0.0050** |
| iliac aneurysm | | uni | 30 (8.2) | 29 (9.3) | 1 (1.9) | 0.89 |
| | | bi | 25 (6.9) | 24 (7.7) | 1 (1.9) | |

**Suppl. Table IIIA: Gender analysis: patient characteristics/metrics and aneurysm characteristics.** Values are given as absolute numbers and percentage and median with 95% confidence interval where applicable. (y=years, CAD=coronary artery disease; COPD=chronic obstructive pulmonary disease; PAOD=peripheral artery occlusive disease; renal insufficiency=serum creatinine >1.2mg/dL; ASS=aspirin; ACE=angiotensin converting enzyme; BMI = body mass index, BSA = body surface area, ASI = aortic size index; CRP=C reactive protein, elevated = >0.5mg/dL; leukocytes elevated = count >10.000/µL, thrombocytes elevated = count > 450.000/µL; p values <0.05 are considered significant and highlighted bold. Chi sqare/Mann Whitney test)

| | | | | total | male | female | p |
|---|---|---|---|---|---|---|---|
| | | | | **364** | 311 | 53 | - |
| **histo-morphology** | | | | | | | |
| HISA | 0 | | | 30 (8.2) | 29 (9.3) | 1 (1.9) | 0.15 |
| | 1 | | | 193 (53.0) | 166 (53.4) | 27 (50.9) | |
| | 2 | | | 107 (29.4) | 89 (24.5) | 18 (34.0) | |
| | 3 | | | 20 (5.5) | 15 (4.8) | 5 (9.4) | |
| AHA | IV | | | 1 (0.3) | 1 (0.3) | 0 | 0.36 |
| | V | | | 69 (19.0) | 59 (19.0) | 10 (18.9) | |
| | VI | | | 217 (59.6) | 182 (25.5) | 35 (66.0) | |
| | VII | | | 19 (5.2) | 17 (5.5) | 2 (3.8) | |
| | VIII | | | 51 (14.0) | 47 (15.1) | 4 (7.5) | |
| media | calcification | | yes | 33 (9.1) | 32 (10.3) | 1 (1.9) | **0.049** |
| | inflammation | Degree | 0 | 204 (56.0) | 175 (56.3) | 29 (54.7) | 0.53 |
| | | | 1 | 132 (36.3) | 114 (36.7) | 18 (34.0) | |
| | | | 2 | 9 (2.5) | 8 (2.6) | 1 (1.9) | |
| | | | 3 | 0 | 0 | 0 | |
| | | Type | no | 205 (56.3) | 176 (56.6) | 29 (54.7) | 0.30 |
| | | | monocytes (C) | 79 (21.7) | 69 (22.2) | 10 (18.9) | |
| | | | granulocytes (A) | 0 | 0 | 0 | |
| | | | plasma cells (C) | 23 (6.3) | 22 (7.1) | 1 (1.9) | |
| | | | mix (A) | 37 (10.2) | 29 (9.3) | 8 (15.1) | |
| | angiogenesis | | yes | 106 (29.1) | 90 (28.9) | 16 (30.2) | 0.85 |
| | elastic fibers | 0 | | 0 | 0 | 0 | 0.52 |
| | | <25 | | 338 (92.9) | 287 (92.3) | 51 (96.2) | |
| | | 25-50 | | 11 (3.0) | 11 (3.5) | 0 | |
| adventitia | inflammation | degree | 0 | 29 (8.0) | 27 (6.8) | 2 (3.8) | 0.15 |
| | | | 1 | 198 (54.4) | 176 (56.6) | 25 (47.2) | |
| | | | 2 | 105 (28.8) | 86 (27.7) | 19 (35.8) | |
| | | | 3 | 19 (5.2) | 14 (4.5) | 5 (9.4) | |
| | | type | no | 29 (8.0) | 27 (8.7) | 2 (3.8) | 0.26 |
| | | | monocytes (C) | 171 (47.0) | 144 (46.3) | 27 (50.9) | |
| | | | granulocytes (A) | 2 (0.5) | 2 (0.6) | 0 | |
| | | | plasma cells (C) | 91 (25.0) | 82 (26.4) | 9 (17.0) | |
| | | | mix (A) | 56 (15.4) | 45 (14.5) | 11 (20.8) | |
| | fibrosis | no | | 1 (0.3) | 0 | 1 (1.9) | 0.53 |
| | | mild | | 109 (29.9) | 97 (31.2) | 12 (22.6) | |
| | | average | | 143 (39.3) | 122 (39.2) | 21 (39.6) | |
| | | severe | | 92 (25.39 | 76 (24.4) | 16 (30.2) | |
| summary | inflammation sum | | 0 | 34 (9.3) | 31 (10.0) | 3 (5.7) | 0.15 |
| | | | 1 | 127 (34.9) | 114 (36.7) | 13 (24.5) | |
| | | | 2 | 133 (36.5) | 106 (34.1) | 27 (50.9) | |
| | | | 3 | 58 (15.9) | 49 (15.8) | 9 (17.0) | |
| | | | 4 | 12 (3.3) | 11 (3.5) | 1 (1.9) | |
| | inflammation type | | 0 | 35 (9.6) | 31 (10.0) | 4 (7.6) | 0.31 |
| | | | A | 63 (17.3) | 50 (16.1) | 13 (24.5) | |
| | | | C | 266 (73.1) | 230 (74.0) | 36 (67.9) | |

**Suppl. Table IIIB: Gender analysis: histo-morphology.** Values are given as absolute numbers and percentage and median with 95% confidence interval where applicable. (HISA = histologic inflammation score aneurysm; AHA = American Heart Association score on atherosclerosis; p values <0.05 are considered significant and highlighted bold. Chi sqare/Mann Whitney test)

| | | total | male | female | P |
|---|---|---|---|---|---|
| | | **364** | 311 | 53 | - |
| **aneurysm growth (n=142: male=119)** | | | | | |
| growth rate (mm/y) | | 3.8 [2.5 – 5.4] | 3.9 [2.5-5.5] | 3.6 [2.8 – 4.3] | 0.97 |
| quart | slow | 36 (25.4) | 31 (26.1) | 5 (21.7) | 0.46 |
| | normal | 70 (49.3) | 56 (47.1) | 14 (60.7) | |
| | fast | 36 (25.4) | 32 (26.9) | 4 (17.4) | |
| **aneurysm morphometry (n=252: male =214)** | | | | | |
| $D_{max}$ (mm) | | 57 [52-69] | 58 [53-70] | 54 [49-60] | **0.005** |
| $Volume_{max}$ ($cm^3$) | | 148 [117 – 201] | 151 [120.8-214] | 115 [83.4-175.1] | 0.12 |
| endosize | α angulation | 18.1 [12.1 – 24.8] | 17.1 [11.8-24.2] | 18.9 [10.4-26.2] | 0.91 |
| | ß angulation | 33.8 [23.3 – 47.0] | 32 [22.5-46.8] | 37.1 [29.2-50] | 0.48 |
| | neck length (mm) | 16 [6 – 33] | 14 [4-31] | 8.5 [3-20] | 0.063 |
| | neck diameter (mm) | 24.0 [21.2 – 26.6] | 25.7 [22.4-28.9] | 22.4 [19.0-25.3] | 0.10 |
| | aortic index | 1.09 [1.06 – 1.14] | 1.08 [1.06-1.13] | 1.12 [1.06-1.19] | 0.50 |
| | iliac index | 1.30 [1.22 – 1.41] | 1.31 [1.24-1.42] | 1.27 [1.18-1.34] | 0.42 |
| **FE analysis (n=206: male=171)** | | | | | |
| local | van Mises stress | 106.2 [84.1 – 132.2] | 109.5 [85.6 – 135.4] | 94.7 [66.4 – 114.2] | **0.01** |
| | PWRI | 0.26 [0.21 – 0.33] | 0.26 [0.21 – 0.32] | 0.3 [0.21 – 0.39] | 0.55 |
| | ILT | 8.2 [3.8 – 15.3] | 7.9 [3.8 – 15.5] | 10.8 [4.2 – 14.0] | 0.48 |
| max | van Mises stress | 205.0 [173.3 – 256.5 | 206.2 [179.2 – 260.0] | 178.0 [149.0 – 211.5] | **0.002** |
| | PWRI | 0.42 [0.34 – 0.56] | 0.42 [0.34 – 0.53] | 0.51 [0.37 – 0.61] | 0.72 |
| | ILT | 19.6 [13.8 – 27.3] | 119.8 [13.9 – 27.9] | 18.0 [13.3 – 22.8] | 0.42 |

**Suppl. Table IIIC: Gender analysis: subgroup analysis.** Values are given as absolute numbers and percentage and median with 95% confidence interval where applicable. (p values <0.05 are considered significant and highlighted bold. Chi sqare/Mann Whitney test)

| | | subgroup | | | | | | | | |
|---|---|---|---|---|---|---|---|---|---|---|
| | | **total** | AAA rupture | p | smoking (current vs. other) | p | diabetes | p | acute (rupt + sympt) | p |
| | | **364** | 52 | | 182 | | 67 | | 89 (3x infl) | |
| **patient characteristics** | | | | | | | | | | |
| male n (%) | | 311 (85.4) | 38 (73.1) | **0.006** | 158 (86.8) | 0.46 | 59 (88.1) | 0.49 | 69 (77.5) | **<0.001** |
| age (y) | | 69 [64-74] | 75 [70-80] | **<0.001** | 68 [62-72] | **<0.001** | 71 [65-75] | 0.42 | 71 [66-77] | **<0.001** |
| comorbidities | hypertension | 295 (81.0) | 38 (86.4)) | 0.51 | 150 (82.4) | 0.59 | 62 (92.5) | **0.009** | 70 (78.7) | 0.46 |
| | diabetes | 67 (18.4) | 8 (18.2) | 0.91 | 35 (19.2) | 0.68 | - | | 17 (19.1) | 0.85 |
| | hyperlipidemia | 199 (54.7) | 13 (29.5) | **<0.001** | 105 (57.7) | 0.25 | 38 (56.7) | 0.69 | 38 (42.7) | **0.009** |
| | CAD | 151 (41.5) | 17 (38.6) | 0.59 | 71 (39.0) | 0.34 | 36 (53.7) | **0.026** | 33 (37.1) | 0.33 |
| | COPD | 69 (19.0) | 12 (27.3) | 0.16 | 44 (24.2) | **0.011** | 13 (19.4) | 0.93 | 20 (22.5) | 0.33 |
| | PAOD | 68 (18.7) | 9 (20.5) | 0.81 | 33 (18.1) | 0.79 | 14 (20.9 | 0.62 | 16 (18.0) | 0.84 |
| | renal insufficiency | 89 (24.5) | 18 (34.6) | 0.065 | 37 (20.3) | **0.018** | 17 (25.4) | 0.86 | 26 (29.2) | 0.23 |
| smoke | current | 182 (50.0) | 15 (60.0) | | | | 35 (52.2) | | 38 (42.7) | |
| | ex | 89 (24.5) | 5 (20.0) | 0.26 | - | | 21 (31.3) | 0.33 | 14 (15.7) | 0.37 |
| | never | 33 (9.1) | 5 (20.0) | | | | 4 (6.0) | | 9 (10.1) | |
| medication | ASS/Clopidogrel | 241 (66.2) | 20 (66.7) | **0.037** | 123 (82.0) | 0.46 | 49 (75.4) | **0.002** | | |
| | ACE inhibitor | 118 (32.4) | 13 (43.3) | **<0.001** | 61 (40.7) | 0.73 | 27 (41.5) | **<0.001** | | |
| | statins | 184 (50.5) | 9 (30) | **<0.001** | 101 (67.3) | 0.06 | 36 (55.4) | **0.004** | - | |
| | metformin | 32 (8.8) | 3 (10) | 0.89 | 11 (7.3) | 0.056 | 30 (46.2) | - | | |
| | insulin | 11 (3.0) | 0 | - | 7 (4.7) | 0.37 | 8 (12.3) | | | |
| **aneurysm characteristics** | | | | | | | | | | |
| ruptured<br>symptomatic<br>asymptomatic<br>inflammatory | | 52 (14.3)<br>37 (10.2)<br>273 (75.0)<br>7 (1.9) | - | | 15 (8.2)<br>23 (12.6)<br>144 (79.1)<br>5 (2.7) | **0.004** | 8 (11.9)<br>9 (13.4)<br>50 (74.6)<br>1 (1.5) | 0.74 | - | |
| $D_{max}$ (mm) | | 57 [52-69] | 77 [62-90] | **<0.001** | 56 [52-65] | 0.05 | 57 [53-65] | 0.60 | 70 [57-82] | **<0.001** |
| iliac aneurysm | uni<br>bi | 30 (8.2)<br>25 (6.9) | 4 (7.7)<br>5 (9.6) | 0.50 | 19 (10.4)<br>14 (7.7) | 0.58 | 3 (4.5)<br>4 (6.0) | 0.51 | 7 (7.9)<br>5 (5.6) | 0.77 |
| **histologic morphology** | | | | | | | | | | |
| angiogenesis (yes) n (%) | | 106 (30.5) | 17 (32.7) | 0.49 | 52 (30.8) | 0.93 | 20 (30.8) | **0**.98 | 26 (30.6) | 0.99 |
| fibrosis | mild | 109 (31.6) | 11 (23.4) | | 44 (25.1) | | 27 (42.9) | | 23 (27.7) | |
| | average | 143 (41.4) | 25 (53.2) | 0.20 | 78 (44.6) | **0.027** | 21 (33.3) | 0.094 | 42 (50.6) | 0.15 |
| | severe | 92 (26.7) | 11 (23.4) | | 53 (30.3) | | 15 (23.8) | | 18 (21.7) | |
| inflammation sum | 0 | 34 (9.3) | 3 (5.8) | | 10 (5.5) | | 5 (7.5) | | 4 (4.5) | |
| | 1 | 127 (34.9) | 18 (34.6) | | 67 (36.8) | | 32 (47.8) | | 28 (31.5) | |
| | 2 | 133 (36.5) | 22 (42.3) | 0.80 | 65 (35.7) | 0.094 | 23 (34.3) | 0.11 | 36 (40.4) | 0.27 |
| | 3 | 58 (15.9) | 7 (13.5) | | 34 (18.7) | | 6 (9.0) | | 18 (20.2) | |
| | 4 | 12 (3.3) | 2 (3.8) | | 6 (3.3) | | 1 (1.5) | | 3 (3.4) | |
| inflammation type | 0 | 35 (9.6) | 3 (5.8) | | 11 (6.0) | | 5 (7.5) | | 4 (4,5) | |
| | acute | 63 (17.3) | 14 (26.9) | 0.11 | 25 (13.3) | **0.007** | 10 (14.9) | 0.65 | 19 (21.3) | 0.11 |
| | chronic | 266 (73.1) | 35 (67.3) | | 146 (80.2) | | 52 (77.6) | | 66 (74.2) | |

**Suppl. Table IV: Additional subgroup analysis.** Values are given as absolute numbers and percentage and median with 95% confidence interval where applicable. (y=years, CAD=coronary artery disease; COPD=chronic obstructive pulmonary disease; PAOD=peripheral artery occlusive disease; renal insufficiency=serum creatinine >1.2mg/dL; ASS=aspirin; ACE=angiotensin converting enzyme; p values <0.05 are considered significant and highlighted bold. Chi sqare/Mann Whitney test)

| | | | subgroup | | |
|---|---|---|---|---|---|
| | | **total** | AAA morphometry | AAA growth | AAA FE analysis |
| | | **364** | 252 | 142 | 206 |
| **patient characteristics** | | | | | |
| male n (%) | | 311 (85.4) | 214 (84.9) | 119 (83.8) | 171 (78.2) |
| age (y) | | 69 [64-74] | 70 [63-75] | 69 [63-74] | 69 [62-74] |
| comorbidities | hypertension | 295 (81.0) | 205 (81.3) | 114 (80.3) | 166 (80.2) |
| | diabetes | 67 (18.4) | 43 (17.1) | 27 (19.0) | 36 (17.5) |
| | hyperlipidemia | 199 (54.7) | 150 (59.5) | 91 (66.2) | 126 (61.2) |
| | CAD | 151 (41.5) | 112 (44.4) | 57 (40.1) | 83 (40.3) |
| | COPD | 69 (19.0) | 51 (20.2) | 28 (19.7) | 39 (18.9) |
| | PAOD | 68 (18.7) | 52 (20.6) | 25 (17.6) | 43 (20.9) |
| | renal insufficiency | 89 (24.5) | 45 (17.9) | 28 (19.7) | 43 (20.9) |
| smoke | current | 182 (50.0) | 136 (54.2) | 73 (51.4) | 112 (54.4) |
| | ex | 89 (24.5) | 61 (24.2) | 41 (28.9) | 52 (25.2) |
| | never | 33 (9.1) | 23 (9.1) | 13 (9.2) | 19 (9.2) |
| medication | ASS/Clopidogrel | 241 (66.2) | 160 (80.8) | 107 (75.4) | 131 (81.4) |
| | ACE inhibitor | 118 (32.4) | 79 (39.9) | 41 (28.9) | 63 (39.1) |
| | statins | 184 (50.5) | 127 (64.1) | 78 (54.9) | 110 (68.3) |
| | metformin | 32 (8.8) | 17 (8.6) | 12 (8.5) | 12 (7.5) |
| | insulin | 11 (3.0) | 8 (4.0) | 2 (1.4) | 7 (4.3) |
| **aneurysm characteristics** | | | | | |
| ruptured | | 52 (14.3) | 35 (13.9) | 10 (7.0) | 23 (11.1) |
| symptomatic | | 37 (10.2) | 28 (11.1) | 9 (6.3) | 21 (10.2) |
| asymptomatic | | 273 (75.0) | 188 (74.6) | 123 (86.6) | 161 (78.2) |
| inflammatory | | 7 (1.9) | 5 (2.0) | 1 (0.7) | 4 (1.9) |
| $D_{max}$ (mm) | | 57 [52-69] | 57 [52-70] | 55 [51-60] | 56 [52-62] |
| iliac aneurysm | uni | 30 (8.2) | 29 (11.5) | 11 (7.7) | 23 (11.2) |
| | bi | 25 (6.9) | 24 (9.5) | 7 (4.9) | 17 (8.3) |
| **histo-morphology** | | | | | |
| angiogenesis (yes) n (%) | | 106 (30.5) | 94 (39.2) | 45 (33.8) | 76 (36.9) |
| fibrosis | mild | 109 (31.6) | 78 (31.6) | 47 (34.6) | 66 (32.0) |
| | average | 143 (41.4) | 110 (44.5) | 52 (38.2) | 87 (42.2) |
| | severe | 92 (26.7) | 56 (22.7) | 35 (25.7) | 47 (22.8) |
| inflammation sum | 0 | 34 (9.3) | 20 (7.9) | 13 (9.2) | 19 (9.2) |
| | 1 | 127 (34.9) | 94 (37.3) | 57 (40.1) | 74 (36.0) |
| | 2 | 133 (36.5) | 96 (38.1) | 48 (33.8) | 81 (39.3) |
| | 3 | 58 (15.9) | 37 (14.7) | 20 (14.1) | 28 (13.6) |
| | 4 | 12 (3.3) | 5 (2.0) | 4 (2.8) | 4 (1.9) |
| inflammation type | 0 | 35 (9.6) | 21 (8.3) | 14 (9.9) | 20 (9.7) |
| | acute | 63 (17.3) | 35 (13.9) | 20 (14.1) | 28 (13.6) |
| | chronic | 266 (73.1) | 196 (77.8) | 108 (76.1) | 158 (76.7) |
| **AAA morphology** | | | | | |
| endosize | α angulation (°) | | 18.1 [12.1 – 24.8] | | |
| | ß angulation (°) | | 33.8 [23.3 – 47.0] | | |
| | neck length (mm) | | 16 [6 – 33] | | |
| | neck diameter (mm) | | 24.0 [21.2 – 26.6] | | |
| | aortic index | | 1.09 [1.06 – 1.14] | | |
| | iliac index | | 1.30 [1.22 – 1.41] | | |
| **AAA growth** | | | | | |
| growth rate (mm/y) | | | | 3.6 [2.5 – 5.3] | |
| quartile | slow | | | 13 (9.1) | |
| | normal | | | 117 (81.8) | |
| | fast | | | 12 (8.4) | |

**Suppl. Table V: Patient cohorts available for subgroup analysis AAA morphometry, growth and FE analyis.** Values are given as absolute numbers and percentage and median with upper/lower quartile range where applicable. (y=years, CAD=coronary artery disease; COPD=chronic obstructive pulmonary disease; PAOD=peripheral artery occlusive disease; renal insufficiency=serum creatinine >1.2mg/dL; ASS=aspirin; ACE=angiotensin converting enzyme)

| | | | | Spearman's Rho | |
|---|---|---|---|---|---|
| | | | | rho | p |
| HISA x inflammation sum (n = 350) | | | | 0.83 | **$3.4e^{-77}$** |
| x inflammation score media (n = 332) | | | | 0.21 | **<0.001** |
| x inflammation score adventitia (n = 341) | | | | 0.92 | **$2.15e^{-142}$** |
| AHA x inflammation sum (n = 345) | | | | -0.1 | 0.06 |
| | | | | p (chi square) | |
| | | acute (n = 60) | chronic (n= 261) | | |
| HISA | 0 | 3 (5.0) | 5 (1.9) | 0.09 | |
| | 1 | 35 (58.3) | 152 (58.2) | | |
| | 2 | 17 (28.3) | 89 (34.1) | | |
| | 3 | 5 (8.3) | 15 (5.7) | | |
| | 4 | - | - | | |

**Suppl. Table VI: HISA/AHA score correlation.** (p values <0.05 are considered significant and highlighted bold. Spearman correlation/Chi sqare test)
